# REGEN-COV Antibody Combination in Outpatients With COVID-19 – Phase 1/2 Results

**DOI:** 10.1101/2021.06.09.21257915

**Authors:** Thomas Norton, Shazia Ali, Sumathi Sivapalasingam, Haitao Gao, Rafia Bhore, Andrea T. Hooper, Jennifer D. Hamilton, Bret J. Musser, Diana Rofail, Joseph Im, Christina Perry, Cynthia Pan, Romana Hosain, Adnan Mahmood, John D. Davis, Kenneth C. Turner, Alina Baum, Christos A. Kyratsous, Yunji Kim, Amanda Cook, Wendy Kampman, Ximena Graber, Gerard Acloque, Yessica Sachdeva, Joseph A. Bocchini, Anita Kohli, Bari Kowal, A. Thomas DiCioccio, Yuhwen Soo, Gregory P. Geba, Neil Stahl, Leah Lipsich, Ned Braunstein, Gary Herman, George D. Yancopoulos, David M. Weinreich, the Trial Investigators

## Abstract

**Background:** Continued SARS-CoV-2 infections and COVID-19-related hospitalizations highlight the need for effective anti-viral treatments in the outpatient setting. In a descriptive interim analysis of the phase 1/2 portion of a double-blind phase 1/2/3 trial in COVID-19 outpatients conducted between June 16, 2020 and September 4, 2020, REGEN-COV^®^ (casirivimab plus imdevimab) antibody combination reduced SARS-CoV-2 viral load versus placebo.

**Methods:** This final phase 1/2 analysis comprises 799 outpatients, including 275 from the previous descriptive analysis (group-1) and 524 from phase 2 (group-2). Patients were randomized (1:1:1) to placebo, REGEN-COV 2400mg, or REGEN-COV 8000mg. Prespecified hierarchical analyses of virologic endpoints were performed in group-2. The proportion of patients with ≥1 COVID-19–related medically attended visit (MAV) through day 29 was assessed in group-1+2. Efficacy was assessed in patients confirmed SARS-CoV-2–positive by baseline nasopharyngeal RT-qPCR. Safety was assessed in all treated patients.

**Results:** Data from 799 outpatients enrolled from June 16, 2020 to September 23, 2020 are reported. Time-weighted average daily reduction in viral load through day 7 was significantly greater in the REGEN-COV combined 2400mg+8000mg group versus placebo in patients with baseline viral load >10^7^ copies/mL (prespecified primary endpoint): -0.68 log_10_ copies/ml (95% CI, -0.94 to -0.41; *P*<.0001). This reduction was - 0.73 (*P*<.0001) and -0.36 (*P*=.0003) log_10_ copies/mL in serum antibody–negative patients and in the overall population, respectively. REGEN-COV reduced the proportion of patients with ≥1 COVID-19–related MAV versus placebo (2.8% [12/434] REGEN-COV combined dose group versus 6.5% [15/231] placebo; *P*=.024; relative risk reduction [RRR]=57%); in patients with ≥1 risk factor for hospitalization, the treatment effect was more pronounced (RRR=71%). Adverse events were similar across groups.

**Conclusions:** In COVID-19 outpatients enrolled prior to the widespread circulation of delta and omicron variants, treatment with REGEN-COV significantly reduced viral load and COVID-19–related MAVs.

## INTRODUCTION

COVID-19 is caused by the severe acute respiratory syndrome coronavirus 2 (SARS-CoV-2), which first emerged in China in December 2019 [1, 2]. COVID-19 severity varies widely among patients, ranging from mild symptoms to severe disease requiring hospitalization or leading to death [3-7]. Higher SARS-CoV-2 viral load is associated with increased disease severity and mortality [8, 9]. Treatments that reduce viral burden may improve COVID-19 outcomes.

REGEN-COV® (casirivimab plus imdevimab; previously REGN-COV2) is a neutralizing monoclonal antibody combination therapy directed against the receptor binding domain (RBD) of the SARS-CoV-2 spike (S) protein [10]. The two human sequence antibodies, casirivimab and imdevimab, bind simultaneously and non-competitively to the RBD of susceptible SARS-CoV-2 variants of concern (VOCs) and, when used in combination, have been shown to protect against viral escape in vitro [11]. In vivo efficacy of REGEN-COV was demonstrated in two animal models (rhesus macaques and golden hamsters), as evidenced by reduced viral load in both models, reduced virus-induced lung pathology in rhesus macaques, and protection from weight loss in golden hamsters [12]. REGEN-COV demonstrates potent in vitro neutralizing activity against SARS-CoV-2 VOCs with increased infectivity (e.g., alpha [B.1.1.7], delta [B.1.617.2]) and those associated with reduced vaccine-induced neutralizing antibody titers or vaccine efficacy (e.g., beta [B.1.351]) but has markedly reduced activity against the omicron BA.1.1 lineage [13-16].

Initial descriptive interim analysis results (N = 275) from the phase 1/2/3 trial assessing the efficacy and safety of REGEN-COV in outpatients with COVID-19 showed that the antibody combination rapidly reduced viral load compared with placebo, with greater reductions in those who at baseline had not yet mounted an endogenous immune response (i.e., were SARS-CoV-2–serum antibody–negative) or had a high viral load [17]. Here, we describe the confirmatory prespecified virologic results from a subsequent phase 2 analysis in 524 additional outpatients with COVID-19, as well as the final phase 1/2 analysis of clinical and safety outcomes in all 799 outpatients.

## METHODS

### TRIAL DESIGN

COV-2067 is an adaptive, multicenter, randomized, double-blind, placebo-controlled, seamless phase 1/2/3 trial in COVID-19 outpatients (NCT04425629) [17, 18].

The design of the phase 1/2 portion of the trial was previously described [17]. Patients were randomly assigned (1:1:1) to receive placebo, 2400 mg REGEN-COV (1200 mg each of casirivimab and imdevimab), or 8000 mg REGEN-COV (4000 mg each of casirivimab and imdevimab) (Figure S1). The phase 2 portion of the trial included a screening/baseline period (days -1 to 1), a follow-up period (days 2 to 25), and an end-of-study visit (day 29). The phase 1 and 2 portions of the trial were identical, except for additional pharmacokinetic analyses and adverse event collection in phase 1.

### INFORMED CONSENT AND TRIAL OVERSIGHT

All patients provided written informed consent before participating in the trial. The trial was conducted in accordance with the principles of the Declaration of Helsinki, GCP/ICH-E-9 guidelines, and all local and international regulatory standards. The local institutional review board or ethics committee at each study center oversaw trial conduct and documentation.

### PATIENTS

Eligible patients were non-hospitalized adults (≥18 years of age) who had a laboratory-confirmed SARS-CoV-2–positive nasopharyngeal (NP) polymerase chain reaction (PCR) test result ≤72 hours prior to randomization and symptoms consistent with COVID-19, as determined by the investigator, with onset ≤7 days before randomization. Patients included in this final phase 1/2 analysis were enrolled in the trial between June 16, 2020, and September 23, 2020. Randomization was stratified by presence or absence of ≥1 risk factor for severe COVID-19, including age ≥50 years, obesity (BMI ≥30 kg/m^2^), immunosuppression, and chronic cardiovascular, metabolic, liver, kidney, or lung disease. The full list of inclusion and exclusion criteria was previously described [17].

### INTERVENTION AND ASSESSMENTS

At baseline (day 1), casirivimab and imdevimab (diluted in a 250-mL normal saline solution for co-administration) or saline placebo was administered intravenously over a period of 1 hour.

All patients were assessed for the presence or absence of anti–SARS-CoV-2 antibodies: anti-spike [S1] IgA, anti-spike [S1] IgG, and anti-nucleocapsid IgG (Euroimmun ELISA assays for anti-S1; Abbott Architect assay for anti-nucleocapsid). As these results were not available at randomization, patients underwent randomization regardless of their baseline serum antibody status and were subsequently grouped for analyses as serum antibody–negative (if all available tests were negative), serum antibody–positive (if any of the tests were positive), or other (missing or inconclusive results).

Central lab detection and quantification of SARS-CoV-2 nucleic acids (viral N protein gene region) utilized a SARS-CoV-2 RT-qPCR swab assay (Viracor Eurofins Clinical Diagnostics).

Additional details on SARS-CoV-2 serology testing and central lab PCR testing were previously described [17]. The protocol contains the full schedule of assessments (Supplementary Appendix).

### ENDPOINTS

The primary virologic endpoint and two key secondary clinical endpoints were prespecified in the phase 1/2 statistical analysis plan. The primary virologic endpoint was defined as the time-weighted average daily change in viral load (log_10_ copies/mL) from baseline (day 1) through day 7. The two key secondary clinical endpoints were (i) the proportion of patients through day 29 with ≥1 medically attended visit (MAV) confirmed by the investigator to be related to COVID-19, where MAV was defined as a hospitalization, emergency room (ER) visit, urgent care visit, or physician office/telemedicine visit, and (ii) the proportion of patients through day 29 with at least 1 MAV consisting of only hospitalizations, ER visits, or urgent care visits.

Safety endpoints for the phase 1/2 portion of the trial included select adverse events, consisting of grade 3 or 4 adverse events (assessed in phase 1 only), serious adverse events (SAEs), and adverse events of special interest (AESIs) defined as grade ≥2 hypersensitivity or infusion-related reactions.

### STATISTICAL ANALYSIS

A previous descriptive analysis reported virologic and clinical efficacy results from patients enrolled in the phase 1/2 portion of the trial from June 16, 2020, to August 13, 2020 (n = 275, analysis group-1) [17]. To confirm the virologic efficacy observed in group-1, analyses of virologic endpoints were conducted using data from patients enrolled in the phase 2 portion from August 14, 2020, to September 23, 2020 (n = 524; analysis group-2). Analyses of clinical endpoints and safety utilized data from patients in both portions (n = 799; group-1+2).

The statistical analysis plan for the presented analysis was finalized prior to database lock and unblinding of the phase 2 dataset. The full analysis set (FAS) included patients with COVID-19 symptoms who underwent randomization. Efficacy analyses of virologic and clinical endpoints were performed in a modified full analysis set (mFAS), comprised of patients with detectable SARS-CoV-2 in baseline NP sample by central lab RT-qPCR (limit of detection, 714 copies/mL). Patients with missing central lab RT-qPCR data were excluded from the mFAS. Subgroup analyses by baseline SARS-CoV-2 serum antibody status and baseline viral load were based on previous descriptive analyses from placebo-treated patients in this study [17] and were prespecified. Safety was assessed in patients in the FAS who received study drug (active or placebo).

Analyses of the primary virologic and two key clinical endpoints were conducted at a two-sided α=0.05 utilizing a hierarchical testing strategy to control for type I error (Table S1). The virologic efficacy endpoint was calculated and analyzed as previously described (see also Supplementary Appendix) [17]. Key secondary clinical endpoints were analyzed using Fisher’s exact test. Statistical analyses were performed with SAS software, version 9.4 or higher (SAS Institute). Additional methods are described in the Supplementary Appendix.

## RESULTS

### BASELINE CHARACTERISTICS

Between June 16, 2020, and September 23, 2020, a time period prior to the emergence of delta and omicron VOCs, 799 symptomatic patients from 54 sites in the United States underwent randomization in the phase 1/2 portion of the trial: 266, 267, and 266 patients were assigned to receive REGEN-COV 2400 mg, REGEN-COV 8000 mg, or placebo, respectively (Figure 1). Among the 799 patients in group-1+2, 87 (10.9%) tested negative for SARS-CoV-2 at baseline via central lab NP RT-qPCR assay and 47 (5.9%) were without central lab baseline viral load data; consequently, the mFAS set comprised 665 patients. Similarly, among the 524 patients in group-2 (primary virologic efficacy analysis), the mFAS set comprised 437 patients.

**Figure 1.**
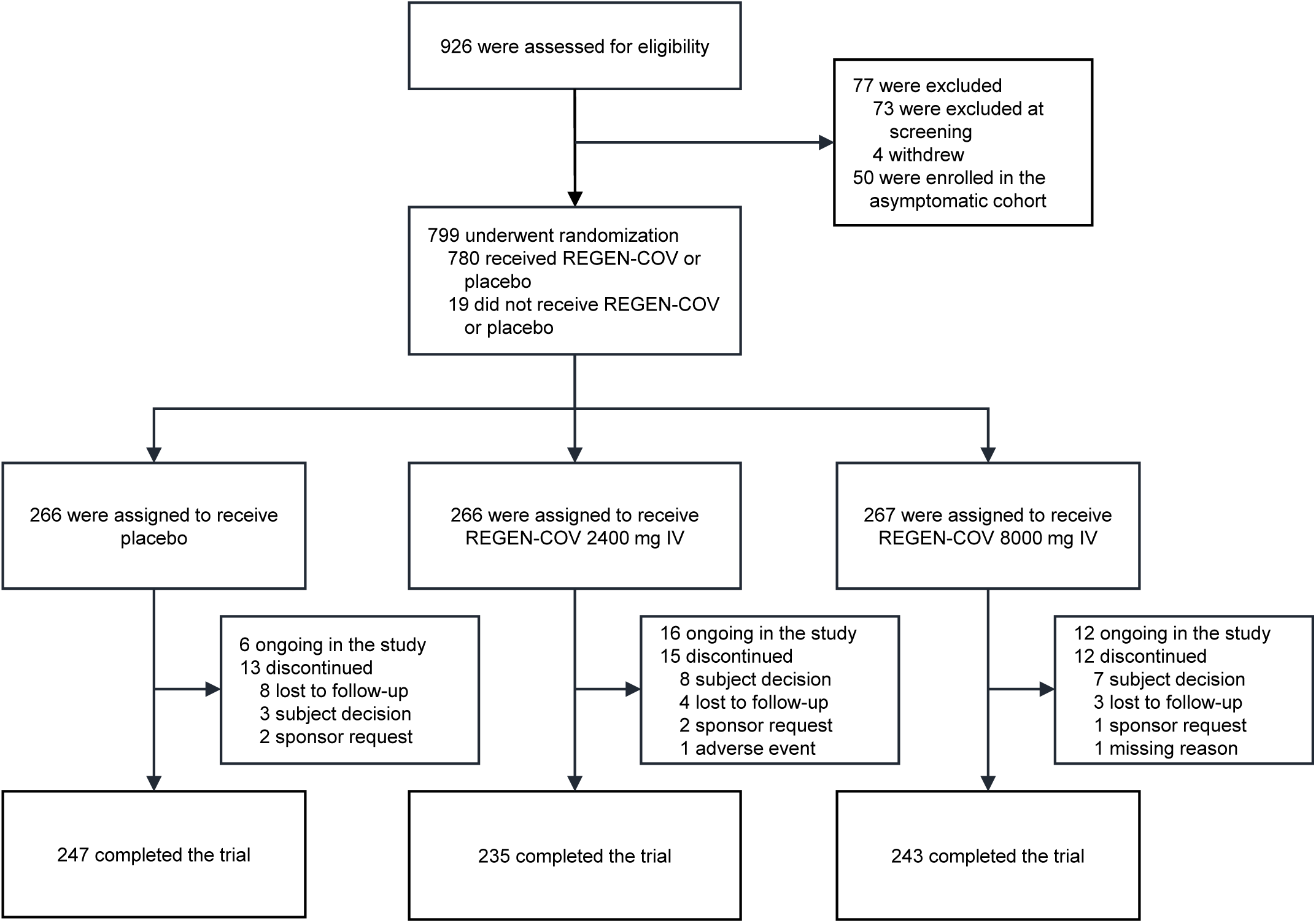
Screening, randomization, and treatment (CONSORT diagram). Abbreviations: IV, intravenous(ly).

Of the 799 randomized patients, the median age was 42.0 years, 47.1% were male, 9.3% identified as Black or African American, and 50.4% identified as Hispanic or Latino (Table 1). Four hundred and eighty-three (60.5%) patients had ≥1 risk factor for hospitalization due to COVID-19, including obesity (BMI ≥30 kg/m^2^; 37.3%), age ≥50 years (29.3%), cardiovascular disease (20.5%), or chronic metabolic disease (13.1%).

**Table 1.** Demographic and Baseline Medical Characteristics (Group-1+2; Full Analysis Set)

| Characteristic | Total<br>(n = 799) | Placebo<br>(n = 266) | REGEN-COV 2400 mg<br>(n = 266) | REGEN-COV 8000 mg<br>(n = 267) | REGEN-COV combined<br>(n = 533) |
| --- | --- | --- | --- | --- | --- |
| Age, y, median<br>(IQR) | 42.0 (31.0–52.0) | 42.0 (32.0–53.0) | 42.0 (31.0–52.0) | 42.0 (30.0–52.0) | 42.0 (30.0–52.0) |
| Male sex | 376 (47.1) | 134 (50.4) | 122 (45.9) | 120 (44.9) | 242 (45.4) |
| Hispanic or<br>Latino ethnic<br>group<sup>a</sup> | 403 (50.4) | 137 (51.5) | 132 (49.6) | 134 (50.2) | 266 (49.9) |
| Race<sup>a</sup> |  |  |  |  |  |
| White | 681 (85.2) | 227 (85.3) | 224 (84.2) | 230 (86.1) | 454 (85.2) |
| Black or African<br>American | 74 (9.3) | 24 (9.0) | 27 (10.2) | 23 (8.6) | 50 (9.4) |
| Asian | 14 (1.8) | 4 (1.5) | 6 (2.3) | 4 (1.5) | 10 (1.9) |
| Native<br>American or<br>Alaska Native | 5 (0.6) | 3 (1.1) | 1 (0.4) | 1 (0.4) | 2 (0.4) |
| Unknown | 7 (0.9) | 3 (1.1) | 1 (0.4) | 3 (1.1) | 4 (0.8) |
| Not reported | 18 (2.3) | 5 (1.9) | 7 (2.6) | 6 (2.2) | 13 (2.4) |
| Weight, kg,<br>median (IQR) | 81.60<br>(69.90–95.30) | 81.80<br>(70.80–94.30) | 81.60<br>(69.90–94.50) | 81.20<br>(68.00–97.10) | 81.60<br>(69.30–95.30) |
| Body-mass index (SD)<sup>b</sup> | 29.67 (8.228) | 29.96 (10.460) | 29.51 (6.413) | 29.57 (7.355) | 29.54 (6.890) |
| Obesity<sup>c</sup> | 298 (37.3) | 93 (35.0) | 101 (38.0) | 104 (39.0) | 205 (38.5) |
| Baseline viral load in nasopharyngeal swab (raw values) |  |  |  |  |  |
| No. of patients | 752 | 259 | 243 | 250 | 493 |
| Viral load, mean, copies/mL (SD) | 18719108.2 (28449769.31) | 20949819.7 (30172071.11) | 17403960.8 (27210806.40) | 17686414.3 (27755446.26) | 17547192.8 (27460771.64) |
| Viral load, median, copies/mL (range) | 304500.0 (1–71000000) | 437000.0 (1–71000000) | 295000.0 (1–71000000) | 262500.0 (1–71000000) | 270000.0 (1–71000000) |
| Baseline viral load in nasopharyngeal swab (log<sub>10</sub> scale) |  |  |  |  |  |
| No. of patients | 752 | 259 | 243 | 250 | 493 |
| Viral load, mean, log<sub>10</sub> copies/mL (SD) | 5.16 (2.500) | 5.21 (2.511) | 5.23 (2.449) | 5.05 (2.544) | 5.14 (2.497) |
| Viral load, median, log<sub>10</sub> copies/mL (range) | 5.48 (0.0–7.9) | 5.64 (0.0–7.9) | 5.47 (0.0–7.9) | 5.42 (0.0–7.9) | 5.43 (0.0–7.9) |
| Baseline viral load in nasopharyngeal swab category |  |  |  |  |  |
| >10<sup>4</sup> | 523 (65.5) | 176 (66.2) | 180 (67.7) | 167 (62.5) | 347 (65.1) |
| >10<sup>5</sup> | 439 (54.9) | 149 (56.0) | 148 (55.6) | 142 (53.2) | 290 (54.4) |
| >10<sup>6</sup> | 332 (41.6) | 114 (42.9) | 110 (41.4) | 108 (40.4) | 218 (40.9) |
| >10<sup>7</sup> | 256 (32.0) | 93 (35.0) | 82 (30.8) | 81 (30.3) | 163 (30.6) |
| Positive baseline qualitative RT-PCR | 665 (83.2) | 231 (86.8) | 215 (80.8) | 219 (82.0) | 434 (81.4) |
| Baseline serum C-reactive protein level |  |  |  |  |  |
| No. of patients | 600 | 203 | 200 | 197 | 397 |
| Level, mean, mg/L (SD) | 13.930 (28.8461) | 19.449 (37.5595) | 10.991 (24.1638) | 11.227 (21.1793) | 11.108 (22.7035) |
| Level, median, mg/L (range) | 3.750 (0.10–239.67) | 4.860 (0.10–232.04) | 3.240 (0.12–239.67) | 3.420 (0.14–157.96) | 3.320 (0.12–239.67) |
| Baseline serum antibody status |  |  |  |  |  |
| Negative | 408 (51.1) | 134 (50.4) | 140 (52.6) | 134 (50.2) | 274 (51.4) |
| Positive | 304 (38.0) | 106 (39.8) | 96 (36.1) | 102 (38.2) | 198 (37.1) |
| Other | 87 (10.9) | 26 (9.8) | 30 (11.3) | 31 (11.6) | 61 (11.4) |
| Symptom onset to randomization |  |  |  |  |  |
| No. of patients | 698 | 240 | 222 | 236 | 458 |
| Time from symptom onset to randomization, median, d (IQR) | 3.0 (2–5) | 3.0 (2–5) | 3.0 (2–5) | 3.0 (2–5) | 3.0 (2–5) |
| At least one risk factor for hospitalization<sup>d</sup> | 483 (60.5) | 158 (59.4) | 165 (62.0) | 160 (59.9) | 325 (61.0) |

Data are presented as no. (%) unless otherwise indicated. Percentages may not total 100 because of rounding.
<sup>a</sup>Race and ethnic group were self-reported by the patients.
<sup>b</sup>The body-mass index is the weight in kilograms divided by the square of the height in meters.
<sup>c</sup>Obesity is defined as a body-mass index of greater than 30.
<sup>d</sup>Risk factors for hospitalization include an age of more than 50 years, obesity, cardiovascular disease (including hypertension), chronic lung disease (including asthma), chronic metabolic disease (including diabetes), chronic kidney disease (including receipt of dialysis), chronic liver disease, and immunocompromised (immunosuppression or receipt of immunosuppressants).
Abbreviations: IQR, interquartile range; RT-PCR, reverse-transcriptase polymerase chain reaction; SD, standard deviation.

A total of 256 (32.0%) patients had baseline viral load >10^7^ copies/mL. The mean time from symptom onset to randomization was 2.9 days in patients with viral load >10^7^ copies/mL and 3.8 days in patients with viral load ≤10^7^ copies/mL, indicating that patients with high viral loads were, on average, earlier in their disease course.

At randomization, 408 (51.1%) patients were SARS-CoV-2 serum antibody–negative, 304 (38.0%) were serum antibody–positive, and 87 (10.9%) were serum antibody– other. Baseline demographic characteristics, including age and the proportion with ≥1 risk factor for hospitalization, were similar between those who were serum antibody– positive compared to those who were serum antibody–negative. Median baseline viral load (log_10_ copies/mL) was 5.48 in the overall trial population, 7.19 serum antibody– negative patients, and 3.68 in serum antibody–positive patients. The mean time from symptom onset to randomization was 3.4 days in the overall trial population, 3.3 days in serum antibody–negative patients, and 3.6 days in serum antibody-positive patients, suggesting that for patients with ≤7 days of symptoms, this measure does not readily discriminate those who have mounted a detectable endogenous immune response from those who have not.

Baseline demographic and clinical characteristics were similar between the 275-patient group-1 and 524-patient group-2 (Table S2) [17] and between the FAS and mFAS populations (Table 1; Table S3).

### NATURAL HISTORY

In the mFAS population, patients in the placebo arm who were serum antibody– negative at baseline had approximately 3-logs higher median viral load at baseline compared with those who were serum antibody-positive (Figure S2) and took substantially longer to bring their viral levels to the lower limit of quantification or to undetectable (Figure S3). Similarly, for COVID-19-related MAVs, placebo-treated patients who were serum antibody–negative at baseline had substantially higher rates of this event (9.7%; 12/124) than placebo-treated patients who were serum antibody– positive at baseline (2.4%; 2/83) (Figure S4). As a detectable endogenous immune response was associated with lower viral titers, there was the expected association of COVID-19–related MAV risk with baseline viral load: MAVs occurred in 0% (0/55) of placebo-treated patients with detectable baseline viral load ≤10^4^ copies/mL versus 8.5% (15/176) with baseline viral load >10^4^ copies/mL. MAVs were also more common in placebo-treated patients with ≥1 risk factor for hospitalization (9.2%; 13/142) vs those with no risk factors (2.2%; 2/89).

### VIROLOGIC EFFICACY

Prespecified comparisons for the virologic efficacy endpoint were assessed hierarchically in the 524-patient group-2 who were confirmed SARS-CoV-2–positive by NP RT-qPCR at baseline (mFAS; n = 437) (Table S1; Table 2). REGEN-COV treatment significantly reduced viral load through day 7 versus placebo in all prespecified virologic efficacy comparisons (Table 2; Figure S5; Figure S6). In the first comparison, among patients with baseline viral load >10^7^ copies/mL, the least-squares mean difference between REGEN-COV treatment (combined 2400 mg and 8000 mg dose groups) and placebo in the time-weighted average (TWA) daily change in viral load through day 7 was -0.68 log_10_ copies/mL (95% CI, -0.94 to -0.41; *P* < .0001) (Table 2; Figure S5). Similarly, the least-squares mean difference in TWA daily change in viral load through day 7 between REGEN-COV treatment and placebo was -0.73 log_10_ copies/mL (95% CI, -0.97 to -0.48; *P* < .0001) in patients who were SARS-CoV-2 serum antibody–negative at baseline, while it was -0.36 log_10_ copies/mL (95% CI, -0.56 to -0.16; *P* = .0003) in the overall population (Table 2; Figure S5). Treatment effects were similar with REGEN-COV 2400 mg and 8000 mg across all virologic efficacy endpoint comparisons (Table 2; Figure S5). Results from additional key virologic endpoints (change from baseline in viral load according to risk factors for hospitalization, time to sustained negative RT-qPCR, and the proportion of patients with high viral load at each visit) are provided in Table S4, Figure S7, and Figure S8.

**Table 2.** Key Virologic and Clinical End Points.

| End Point | Placebo | REGEN-COV 2400 mg | REGEN-COV 8000 mg | REGEN-COV combined |
| --- | --- | --- | --- | --- |
| **Time-weighted average change from baseline in viral load<sup>a</sup> (log<sub>10</sub> copies/mL) from day 1 through day 7 (analysis group 2)** |  |  |  |  |
| **Baseline viral load >10<sup>7</sup> copies/mL (mFAS)** |  |  |  |  |
| No. of patients | 70 | 58 | 52 | 110 |
| Least-squares mean change, log<sub>10</sub> copies/mL (SE) | -1.46 (0.15) | -2.14 (0.16) | -2.13 (0.16) | -2.13 (0.13) |
| 95% CI | -1.75, -1.16 | -2.44, -1.83 | -2.44, -1.82 | -2.39, -1.87 |
| Difference vs placebo at day 7 — log<sub>10</sub> copies/mL |  |  |  |  |
| Least-squares mean (SE) |  | -0.68 (0.16) | -0.68 (0.16) | -0.68 (0.14) |
| 95% CI<sup>b</sup> (P value) |  | -0.99, -0.37 (<.0001) | -0.99, -0.36 (<.0001) | -0.94, -0.41 (<.0001) |
| **Baseline viral load >10<sup>6</sup> copies/mL (mFAS)** |  |  |  |  |
| No. of patients | 86 | 72 | 73 | 145 |
| Least-squares mean change, log<sub>10</sub> copies/mL (SE) | -1.40 (0.13) | -2.13 (0.13) | -1.98 (0.13) | -2.06 (0.11) |
| 95% CI | -1.65, -1.14 | -2.38, -1.88 | -2.24, -1.72 | -2.27, -1.85 |
| Difference vs placebo at day 7 — log<sub>10</sub> copies/mL |  |  |  |  |
| Least-squares mean (SE) |  | -0.73 (0.14) | -0.58 (0.14) | -0.65 (0.12) |
| 95% CI<sup>b</sup> (P value) |  | -1.01, -0.45 (<.0001) | -0.86, -0.30 (<.0001) | -0.89, -0.41 (<.0001) |
| **Baseline serum antibody status: negative (mFAS)** |  |  |  |  |
| No. of patients | 93 | 80 | 77 | 157 |
| Least-squares mean change, log<sub>10</sub> copies/mL (SE) | -1.18 (0.10) | -1.92 (0.11) | -1.90 (0.11) | -1.91 (0.08) |
| 95% CI | -1.38, -0.99 | -2.13, -1.71 | -2.11, -1.69 | -2.06, -1.76 |
| Difference vs placebo at day 7 — log<sub>10</sub> copies/mL |  |  |  |  |
| Least-squares mean (SE) |  | -0.74 (0.14) | -0.71 (0.14) | -0.73 (0.12) |
| 95% CI<sup>b</sup> (P value) |  | -1.02, -0.45 (<.0001) | -1.00, -0.43 (<.0001) | -0.97, -0.48 (<.0001) |
| **mFAS** |  |  |  |  |
| No. of patients | 146 | 137 | 141 | 278 |
| Least-squares mean change, log<sub>10</sub> copies/mL (SE) | -1.30 (0.09) | -1.69 (0.09) | -1.64 (0.09) | -1.66 (0.07) |
| 95% CI | -1.49, -1.12 | -1.87, -1.50 | -1.82, -1.46 | -1.81, -1.52 |
| Difference vs placebo at day 7 — log<sub>10</sub> copies/mL |  |  |  |  |
| Least-squares mean (SE) |  | -0.38 (0.12) | -0.34 (0.12) | -0.36 (0.10) |
| 95% CI<sup>b</sup> (P value) |  | -0.61, -0.15 (.0011) | -0.57, -0.11 (.0035) | -0.56, -0.16 (.0003) |
| **Proportion of patients with COVID-19–related MAVs through day 29 (analysis groups 1+2)** |  |  |  |  |
| **mFAS** |  |  |  |  |
| No. of patients | 231 | 215 | 219 | 434 |
| Patients with ≥1 visit within 29 days | 15 (6.5) | 6 (2.8) | 6 (2.7) | 12 (2.8) |
| Difference vs placebo |  | -3.7% | -3.8% | -3.7% |
| 95% CI<sup>b</sup> (P value) |  | -8.0, 0.3 (.0754) | -8.1, 0.2 (.0737) | -7.9, -0.3 (.0240) |
| **Proportion of patients with one of a subset of COVID-19–related MAVs consisting of hospitalization, ER visit, or urgent care visit through day 29 (analysis groups 1+2)** |  |  |  |  |
| **mFAS** |  |  |  |  |
| No. of patients | 231 | 215 | 219 | 434 |
| Patients with ≥1 visit within 29 days | 10 (4.3) | 5 (2.3) | 5 (2.3) | 10 (2.3) |
| Difference vs placebo |  | -2.0% | -2.0% | -2.0% |
| 95% CI<sup>b</sup> (*P* value) |  | -11.2, 7.3 (.2983) | -11.3, 7.2 (.2962) | -10.0, 6.0 (.1575) |

Data are presented as no. (%) unless otherwise indicated.
<sup>a</sup>The time-weighted mean change in viral load was based on an analysis of covariance model with treatment group, risk factor, and baseline antibody status as fixed effects and baseline viral load and treatment group-by-baseline viral load as covariates. Confidence intervals were not adjusted for multiplicity.
<sup>b</sup>Confidence intervals for the difference (REGEN-COV minus placebo) were based on the exact method and were not adjusted for multiplicity. Abbreviations: CI, confidence interval; mFAS, modified full analysis set; SE, standard error.

### CLINICAL EFFICACY

There were two clinical efficacy endpoints prespecified for hierarchical testing: the proportion of patients with ≥1 COVID-19–related MAV and the proportion of patients with at least one of a subset of COVID-19–related MAVs, consisting of hospitalizations, ER visits, or urgent care visits (Table S1; Table 2). Both endpoints were assessed through day 29 in patients from the pooled 799-patient group (group-1+2) who were confirmed SARS-CoV-2–positive by NP RT-qPCR at baseline (mFAS; n = 665). Overall, 67% of the COVID-19–related MAVs were hospitalizations or ER visits (30% and 37%, respectively), 26% physician office/telemedicine visits, and 7% urgent care visits. Descriptions of COVID-19–related MAVs are included in Table S5.

The proportion of patients in the REGEN-COV treatment group (combined 2400 mg and 8000 mg dose groups) with ≥1 COVID-19–related MAV was 2.8% (12 of 434) compared with 6.5% (15 of 231) in the placebo group, which represents a relative reduction of 57% (absolute difference vs placebo, -3.7 percentage points; 95% CI, -8% to 0%; *P* = .024) (Table 2). The proportion of patients with ≥1 COVID-19–related hospitalization, ER visit, or urgent care visit was numerically lower in the REGEN-COV combined dose group (2.3%) vs placebo (4.3%), but the difference did not reach statistical significance (Table 2). For these two clinical endpoints, a similar benefit with REGEN-COV was also observed when analyses were conducted based on the FAS (Table S6). Post-hoc analyses showed that REGEN-COV (combined dose group) reduced the proportion of patients with ≥1 COVID-19–related hospitalization (0.7% vs 2.2% placebo; 68% relative reduction) and the proportion of patients with ≥1 COVID-19–related hospitalization or ER visit (1.8% vs 4.3% placebo; 57% relative reduction) (Table S7).

Additional post hoc analyses investigated the effects of the antibody combination treatment on MAVs in various subgroups. For patients who were serum antibody-negative at baseline, REGEN-COV (combined dose group) reduced the proportion of patients with ≥1 COVID-19–related MAV compared with placebo (3.4% vs 9.7%; 65% relative reduction) (Table S8). The treatment effect was even more pronounced in patients with ≥1 risk factor for severe disease (n = 408), where REGEN-COV (combined dose group) reduced the proportion of patients with ≥1 COVID-19–related MAV versus placebo (2.6% vs 9.2%; 71% relative reduction) (Figure 2; Table S9). For patients with ≥1 risk factor who, at baseline, were also serum antibody–negative *and* had a viral load >10^4^ copies/mL (n = 217), REGEN-COV (combined dose group) led to an 84% reduction in the proportion of patients with ≥1 COVID-19–related MAV compared with placebo: 2.1% vs 13.2% (Table S10).

**Figure 2.**
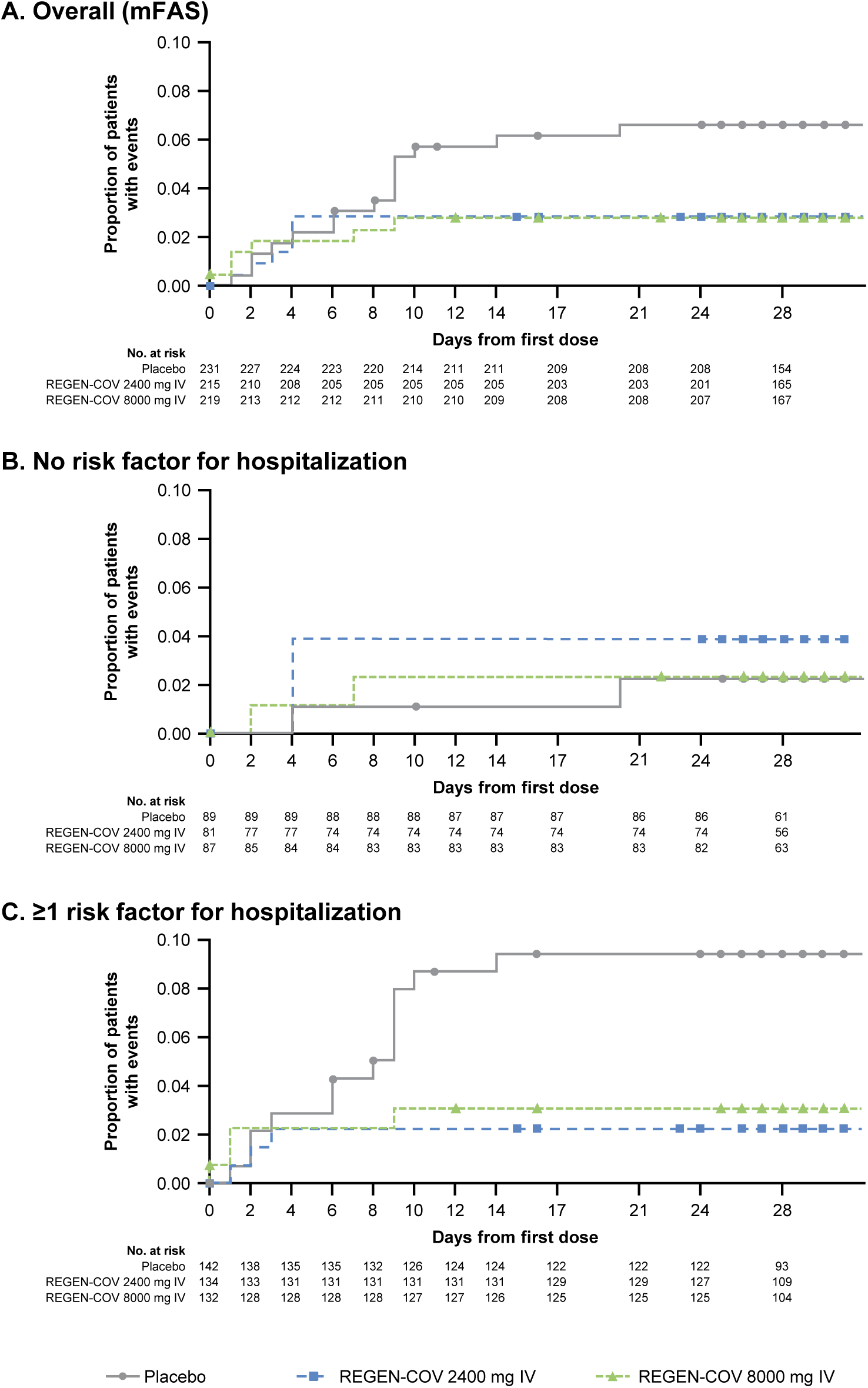
Proportion of patients with ≥1 COVID-19–related medically attended visit (MAV) (A) in the overall population (modified full analysis set [mFAS]), (B) with no risk factor for hospitalization, and (C) with ≥1 risk factor for hospitalization. MAV was defined as a hospitalization or ER, urgent care, or physician office/telemedicine visit that was confirmed by the investigator to be related to COVID-19. Abbreviations: IV, intravenous(ly).

The majority (59%) of patients who experienced a MAV had a viral load of ≥4 log_10_ copies/mL around the time of the MAV (Table S5; Figure S9). As with the virologic endpoints, no meaningful differences in clinical outcomes were observed between low-dose and high-dose treatments.

### SAFETY

No patients died in the phase 1/2 portion of the study. SAEs were experienced by 4 of 258 patients (1.6%) in the REGEN-COV 2400-mg group, 2 of 260 patients (0.8%) in the REGEN-COV 8000-mg group, and 6 of 262 patients [2.3%] in the placebo group (Table 3; Table S11). All SAEs were considered to be due to advanced or progressive COVID-19 disease and/or associated concomitant clinical conditions and were not evaluated to be related to the study drug treatment.

**Table 3.** Overview of Serious Adverse Events and Adverse Events of Special Interest in the Safety Population.

| Event | Placebo<br>(n = 262) | REGEN-COV 2400 mg<br>(n = 258) | REGEN-COV 8000 mg<br>(n = 260) | REGEN-COV combined<br>(n = 518) |
| --- | --- | --- | --- | --- |
| Any serious adverse event | 6 (2.3) | 4 (1.6) | 2 (0.8) | 6 (1.2) |
| Any adverse event of special interest<sup>a</sup> | 2 (0.8) | 0 | 4 (1.5) | 4 (0.8) |
| Any serious adverse event of special interest<sup>a</sup> | 0 | 0 | 0 | 0 |
| Grade ≥2 infusion-related reaction within 4 days | 1 (0.4) | 0 | 4 (1.5) | 4 (0.8) |
| Grade ≥2 hypersensitivity reaction within 29 days | 2 (0.8) | 0 | 0 | 0 |
| Patients with adverse events that occurred or worsened during the observation period<sup>b</sup> |  |  |  |  |
| Patients with grade 3 or 4 event | 4 (1.5) | 3 (1.2) | 2 (0.8) | 5 (1.0) |
| Patients with adverse event that led to death | 0 | 0 | 0 | 0 |
| Patients with adverse event that led to withdrawal from the trial | 0 | 0 | 1 (0.4) | 1 (0.2) |
| Patients with adverse event that led to infusion interruption* | 1 (0.4) | 0 | 1 (0.4) | 1 (0.2) |

Data are presented as no. (%).
<sup>a</sup>Events were grade 2 or higher hypersensitivity reactions or infusion-related reactions.
<sup>b</sup>Events listed here were not present at baseline or were an exacerbation of a preexisting condition that occurred during the observation period, which is defined as the time from administration of REGEN-COV or placebo to the final follow-up visit.

AESIs (grade ≥2, infusion-related reactions and hypersensitivity reactions) that occurred or worsened during the safety observation period were reported in no patients in the 2400-mg group, 4 (1.5%) patients in the 8000-mg group, and 2 (0.8%) patients in the placebo group (Table 3; Table S11).

### PHARMACOKINETICS

The mean concentrations for casirivimab and imdevimab increased in a dose-proportional manner and were consistent with linear pharmacokinetics for single intravenous doses (Table S12). The mean±SD day 29 concentrations of casirivimab and imdevimab in serum were 79.7±34.6 and 65.2±28.1 mg/L, respectively, for the low (1200 mg) doses and 250±97.4 and 205±82.7 mg/L, respectively, for the high (4000mg) doses (Table S12).

## DISCUSSION

The findings from this final phase 1/2 analysis of REGEN-COV antibody combination for the treatment of outpatients with COVID-19 confirm and extend the findings from the first 275 patients [17]. To better understand the natural history of COVID-19 in outpatients, data from placebo patients in this trial were examined. These data confirm previous findings that patients who, at baseline, had not yet mounted their own immune response (i.e., serum antibody–negative) had baseline median viral loads that were almost 3 log copies/mL higher compared with patients who were serum antibody– positive and took longer to reach low or undetectable levels [17, 19, 20]. Similar to other viral infections, such as HIV [21], Ebola virus [22], and influenza [23], high viral load appears to be a predictor of disease progression in COVID-19, as evidenced by the fact that, in placebo-treated patients, COVID-19–related MAVs exclusively occurred in patients with baseline viral loads >10^4^ copies/mL. The data also indicate that risk factors for severe disease, such as older age and obesity, may help to predict outpatients who are most likely to have a subsequent COVID-19–related MAV. For example, 9.2% (13/142) of placebo patients with ≥1 risk factor had a MAV compared with 2.2% (2/89) of placebo patients without any risk factors. In this trial, >80% of patients with risk factors were serum antibody–negative or had a viral load >10^4^ copies/mL. In the absence of a rapid serology test or quantitative PCR assay to identify at-risk patients, those with risk factors for hospitalization should be strongly considered for early treatment with the antibody combination.

The prespecified hierarchical analysis described herein prospectively, and with high statistical significance, replicates and confirms the virologic efficacy of REGEN-COV as previously reported in an earlier descriptive analysis and reveals similar virologic efficacy with both the 2400-mg and 8000-mg doses of the antibody combination [17]. The reduction in viral load was greatest in the first 5 days after treatment and was most pronounced in patients who, at baseline, were SARS-CoV-2 serum antibody–negative or who had high viral load.

The reduction in viral load after treatment with either dose of REGEN-COV was accompanied by a significant reduction in the proportion of patients requiring a subsequent COVID-19–related MAV, the majority (67%) of which were hospitalizations or ER visits. REGEN-COV treatment led to a 57% relative reduction in MAVs (6.5% placebo vs 2.8% combined dose group; *P* = .0240). Interestingly, the reduction in the proportion of patients with MAVs treated with REGEN-COV compared with placebo became evident after the first week of treatment. One possible explanation for this finding is that the occurrence of early MAVs may be less modifiable despite accelerated clearance of the virus. Indeed, among patients treated with REGEN-COV, all 3 hospitalizations occurred in the first 3 days after treatment when viral loads were still ≥4 log_10_ copies/mL, while no hospitalizations occurred after day 7 (Table S5; Figure S9). In contrast, among patients treated with placebo, 3 of the 5 hospitalizations occurred after day 7, when viral loads continued to be high (≥4 log_10_ copies/mL). These data support early identification and rapid treatment of outpatients with COVID-19 in order to optimize the efficacy of REGEN-COV treatment.

The safety of REGEN-COV antibody combination was as previously reported [17]. Low incidences of SAEs, infusion-related reactions, and hypersensitivity reactions were observed. There were no adverse findings observed in patients who were serum antibody–positive at baseline. Similar to results reported previously [17], concentrations of each antibody in serum at day 29 were well above the predicted neutralization target concentration based on in vitro and preclinical data [10-12].

The clinical evidence from the phase 1/2 portion of this study, conducted between June and October 2020, suggests that treatment with REGEN-COV has the greatest benefit when given to high-risk patients who present early after diagnosis when they are most likely to have high viral load. Based on these results, REGEN-COV 2400 mg received Emergency Use Authorization (EUA) from the US FDA in November 2020 for the treatment of mild-to-moderate COVID-19 in patients at high risk for progressing to severe COVID-19 (note: REGEN-COV 1200 mg was subsequently authorized based on phase 3 data) [18, 24]. In January 2022, the FDA amended the EUA for REGEN-COV to exclude its use in geographic regions where infection or exposure is likely to have been caused by a non-susceptible SARS-CoV-2 variant, such as the omicron BA.1.1 sublineage [25].

In this pandemic, early treatment of COVID-19 outpatients is crucial and, if unable to rapidly determine viral load or serum antibody status, the risk-benefit assessment supports treatment to prevent MAVs in high-risk patients.

## Supporting information

AB ICMJE

AC ICMJE

AH ICMJE

AK ICMJE

AM ICMJE

BK ICMJE

BM ICMJE

CK ICMJE

CPan ICMJE

CPerry ICMJE

DR ICMJE

DW ICMJE

GH ICMJE

GPG ICMJE

GY ICMJE

HG ICMJE

JB ICMJE

JD ICMJE

JH ICMJE

JI ICMJE

KT ICMJE

LL ICMJE

NB ICMJE

NS ICMJE

RB ICMJE

RH ICMJE

SA ICMJE

SS ICMJE

TD ICMJE

TN ICMJE

WK ICMJE

XG ICMJE

YK ICMJE

YSachdeva ICMJE

YSoo ICMJE

CONSORT checklist

## Data Availability

Qualified researchers may request access to study documents (including the clinical study report, study protocol with any amendments, blank case report form, statistical analysis plan) that support the methods and findings reported in this manuscript. Individual anonymized participant data will be considered for sharing once the indication has been approved by a regulatory body, if there is legal authority to share the data and there is not a reasonable likelihood of participant re-identification. Submit requests to https://vivli.org/.

## DATA SHARING

A data sharing statement provided by the authors is available with the full text of this article.

## SUPPORTED BY

Supported by Regeneron Pharmaceuticals, Inc. Certain aspects of this project have been funded in whole or in part with federal funds from the Department of Health and Human Services; Office of the Assistant Secretary for Preparedness and Response; Biomedical Advanced Research and Development Authority, under OT number: HHSO100201700020C.

## ROLE OF THE FUNDER/SPONSOR

This work was supported by Regeneron Pharmaceuticals, Inc. and the Biomedical and Advanced Research and Development Authority of the Department of Health and Human Services (ClinicalTrials.gov number, NCT04425629). Regeneron designed the trial and, with the trial investigators, gathered the data. Regeneron analyzed the data. The investigators, site personnel, and Regeneron were unaware of the treatment-group assignments. An independent data and safety monitoring committee monitored unblinded data to make recommendations about trial modification and termination.

Regeneron and all authors were responsible for preparation, review, and approval of the manuscript. Regeneron did not have the right to veto publication or to control the decision to which journal the paper was submitted. All final content decisions were made by the authors.

## ACKNOWLEDGEMENT

We thank the study participants; their families; the investigational site members involved in this trial (principal and subprincipal investigators, listed in the Supplementary Appendix); the Regeneron trial team (members listed in the Supplementary Appendix); the members of the independent data and safety monitoring committee; Brian Head, Ph.D., Caryn Trbovic, Ph.D., and S. Balachandra Dass, Ph.D., from Regeneron Pharmaceuticals for assistance with development of an earlier version of the manuscript; and Prime for formatting and copy editing suggestions for an earlier version of the manuscript.

