## Supplementary Appendix for "REGEN-COV Antibody Combination in Outpatients With COVID-19 – Phase 1/2 Results"

#### Study Sites and Investigators

**AGA Clinical Trials, Miami, FL:** Dario Altamirano, Faisal Fakih, Dickson Ellington

**Arizona Liver Health, Mesa, AZ:** Yessica Sachdeva, Anita Kohli, Amanda McFarland, Dina Gibson

**Arizona Liver Health, Tucson, AZ:** Anita Kohli, Vicki McIntyre, Yessica Sachdeva

**Ark Clinical Research, Long Beach, CA:** Jason Ahn, Kenneth Kim, Lisa Neinchel, Nayna Paryani, Amber Mottola, Eva Day, Martha Navarro

**Atella Clinical Research, La Palma, CA:** Rafaelito Victoria, Xanthe Victoria, Rene Uong

**Avera McKennan Hospital and University Health Center, Sioux Falls, SD:** Jawad Nazir, John Lee, Amy Elliott, Touba Naim, Khizar Hamid, Muhammad Hamza, Robert Kessler, Kara Bruning

**Baylor University Medical Center, Dallas, TX:** Mezgebe Berhe, Haley Clinton, Uriel Sandkovsky, Emma Dishner, Rahaf Al Masri, Erin Duhaime, Christopher Bettacchi

**Bio-Medical Research, LLC, Miami, FL:** Lilia Roque-Guerrero, Ana Gomez Ramirez, Javier Capote, Gisel Paz

**Carolina Medical Research, Clinton, SC:** Nancy Patel, Ravikumar Patel, Ryan Sattar

**Catalina Research Institute, Montclair, CA:** Rizwana Mohseni, Shelia De Jesus-Maranan, Cecilia Casaclang

**Clinical Research of Central Florida, Winter Haven, FL:** Robinson Koilpillai, Stephanie Cassady, Jennifer Cox, Eduardo Torres

**Crossroads Clinical Research, Corpus Christi, TX:** Michael Winnie, Jerry Plemons, Omesh Verma, Richard Leggett

**Epic Medical Research, Red Oak, TX:** Haresh Boghara, Sunny Patel, Bari Eichelbaum

**Florida Pulmonary Research Institute, LLC, Winter Park, FL:** Faisal A. Fakih, Faisal M. Fakih, Fernando Alvarado, Daniel Layish, Jose Diaz, Andres Perez

**Fomat Medical Research, Oxnard, CA:** Augusto Focil, Griselda Rosas, Stevan Correa, Michael Bogseth

**Global Clinical Professionals Research, Saint Petersburg, FL:** Roxana Stoici, Gualberto Perez, Joseph Pica, Enrique Villareal

**Holy Name Medical Center, Teaneck, NJ:** Suraj Saggar, Thomas Birch, Benjamin De La Rosa, Karyna Neyra, Erina Kunwar

**Hope Clinical Research, Canoga Park, CA:** Hessam Aazami, Cheryl Bland, Meera Patel, Jamsheed Akhavan

**IACT Health, Columbus, GA:** Jeffrey Kingsley, April Pixler

**Inova Health Care Services (INOVA Fairfax Hospital), Falls Church, VA:** Christopher deFilippi, Steven Nathan, Lindsay Clevenger

**Lincoln Medical Center - NYC Health + Hospitals, Bronx, NY:** Vidya Menon, Moiz Kasubhai, Usha Venugopal, Anjana Pillai, Franscene Oulds

**Long Beach Clinical Trials, Long Beach, CA:** Barry Heller, Monica Lee, Alvin Merilles, Deirdre Heimer

**Long Beach Medical Center, Long Beach, CA:** Jimmy Johannes, Thomas Jaing, Christopher Yee, Henry Su, Andrew Wittenberg, Anthony Arguija

**Medical University of South Carolina, Charleston, SC:** Eric Meissner, Patrick Flume, Andrew Goodwin, Deeksha Jandhyala, Nandita Nadig

**Mercury Clinical Research, Houston, TX:** Rajasekaran Annamalai, Huy Nguyen, Nizar Nayani, Mahalakshmi Ramchandra

**Midland Florida Clinical Research Center, Deland, FL:** Godson Oguchi, DeAndrea Duffus

**Midway Immunology and Research Center, Fort Pierce, FL:** Moti Ramgopal, Brenda Jacobs, Lisa Cason, Angela Trodglen

**Next Level Urgent Care, Houston, TX:** Terence Chang, Robbyn Traylor, Lenee Gordon, John McDivitt, Lizette Castro

**PharmaTex Research, Amarillo, TX:** David Brabham, Mark Sigler, Tarek Naguib

**PMG Research of McFarland Clinic, Ames, IA:** Jennifer Killion, Rupal Amin, Shauna Basener, Timothy Lowry

**PMG Research of Wilmington, Wilmington, NC:** Kevin Cannon, Mesha Chadwick

**Providence Saint John's Health Center, Santa Monica, CA:** Terese Hammond, Fabian Andres Romero, Steven O'Day, Trevan Fischer, Ana Rocha, Anmol Rangoola

**Queens NYC Health + Hospitals, Jamaica, NY:** Jazila Mantis, Margaret Kemeny, Merjona Saliaj

**Qway, Hialeah, FL:** Oscar Galvez, Fausto Castillo

**Remington-Davis, Columbus, OH:** Edward Cordasco, Brian Zeno, Heather Holmes, Heather Lee

**Ruane Clinical Research Group, Los Angeles, CA:** Peter Ruane, Peter Wolfe, Kenny Trinidad, Isaac Berlin

**Sarasota Memorial Hospital, Sarasota, FL:** Manuel Gordillo, Rishi Bhattacharyya, Sudha Tallapragada, Annette Artau, Julie Larkin, Roberto Mercado, Michael Milam, Natan Kraitman, Sarah Temple, Lenka Offner, Rabih Loutfi, Kirk Voelker, Michael Lowry, Marshall Frank, Ashley Grant

**Stanford University, Palo Alto, CA:** Upinder Singh, Aruna Subramanian, Yvonne Maldonado, Jason Andrews, Chaitan Khosla

**Sun Research Institute, San Antonio, TX:** Carl Dukes, Robert Bass, Larry Lothringer, Leonel Reyes

**Tandem Clinical Research, Maitland, FL:** Esteban Olivera, Maya Abreu

**Tandem Clinical Research, Marrero, LA:** Adil Fatakia, Marissa Miller, Kristen Clinton, Gary Reiss

**Temple University Hospital (TUH), Philadelphia, PA:** Gerard Criner, Nathaniel Marchetti, Parag Desai, Daniel Salerno, Fredric Jaffe, Samuel Krachman, Matthew Zheng, Maulin Patel, Junad Chowdhury, Daniel Mueller

**The George Washington University Hospital, Washington, DC:** David Diemert, Afsoon Roberts, David Parenti, Hana Akselrod, Marc Siegel, Andrew Meltzer, Elissa Malkin

**The University of Texas Health Science Center, Tyler, TX:** Julie Philley, Megan Devine, Richard Yates, Steven Hickerson

**Triple O Research Institute PA, West Palm Beach, FL:** Olayemi Osiyemi, Jose A. Menajovsky-Chaves, Christina Campbell

**Universal Medical and Research Center, LLC, Miami, FL:** Gerard Acloque, Agustin Martinez

**University of Colorado, Aurora, CO:** Thomas Campbell, Martin Krsak, Steven Johnson, Hillary Dunlevy

**University of Iowa, Iowa City, IA:** Alejandro Comellas, Joel Kline, Spyridon Fortis

**University of South Florida, Tampa, FL:** Kami Kim, Seetha Lakshmi, Tiffany Vasey, Asa Oxner, Jason Wilson, Lucy Guerra

**University of Texas (UT) - Southwestern Medical Center, Dallas, TX:** Satish Mocherla, Mamta Jain, Jessica Meisner, Nancy Rollins

**University of Wisconsin, Madison, WI:** William Hartman, Joseph Connor, Richard Striker

**Willis-Knighton Physician Network, Shreveport, LA:** Joseph Bocchini, Clint Wilson

**Xera Med Research, Boca Raton, FL:** Anna Martin, Gargi Gharat, Candace Kokaram, Ket Wray, Clement Partap, Ulyana Arzamasova, Kristina Louissaint, Maria Fernandez

**Xera Med Research, Miami, FL:** Anna Martin, Ket Wray, Kristina Louissaint, Maria Fernandez, Gargi Gharat

#### Regeneron Study Team

Achint Chani, Adebiyi Adepoju, Adnan Mahmood, Aisha Mortagy, Ajla Dupljak, Alison Brown, Alpana Waldron, Amanda Cook, Amy Froment, Andrea Hooper, Andrea Margiotta, Andrew Bombardier, Anne Smith, Aswani Bathula, Bari Kowal, Barry Siliverstein, Benjamin Horel, Bret Musser, Brian Bush, Brian Head, Bryan Zhu, Camille Debray, Careta Phillips, Carol Lee, Caryn Trbovic, Catherine Elliott, Chad Fish, Charlie Ni, Charlotte Lyon, Christina Perry, Christine Enciso, Christopher Caira, Christopher Chamak, Christopher Powell, Cliff Baum, Colby Burk, Cynthia Pan, David Liu, David Stein, Daya Gulabani, Deborah Leonard, Denise Bonhomme, Denise Kennedy, Derrick Bramble, Dhanalakshmi Barron, Diana Rofail, Dipinder Kaur, Dona Bianco, Donna Gambaccini, Eduardo Forleo Neto, Edward Jean-Baptiste, Ehsan Bukhari, Elizabeth Bucknam, Emily Nanna, Esther Huffman O'Keefe, Evelyn Gasparino, Georgia Bellingham, Giane Sumner, Grainne Moggan, Grainne Power, Haitao Gao, Haixia Zeng, Heath Gonzalez, Helen Kang, Hibo Noor, Ian Minns, James Donohue, Janice Austin, Janie Parrino, Jeannie Yo, Jenna McDonnell, Jennifer Hamilton, Jessica Boarder, Jing Xiao, Jingchun Yu, Joanne Malia, Joanne Tucciarone, John Strein, Jonathan Cohen, Jordan Ursino, Joseph Im, Joseph Wolken, Karen Browning, Karen Yau, Kenneth Turner, Kimberly Dornheim, Kit Chiu, Kristina McGuire, Kristy Macci, Kurt Ringleben, Kyle Foster, Lacey Douthat, Latora Knighton, Lisa Boersma, Lisa Hersh, Lisa Purcell, Lisa Sherpinsky, Lori Geissler, Marc Dickens, Marco Mancini, Martha Simpkins, Meagan O'Brien, Michael Batchelder, Michael Partridge, Michal Rozanski, Michel Tarabocchia, Michelle Wong, Mivianisse Rodriguez, Moetaz Albizem, Muriel O'Byrne, Nagendher Burra, Neena Sarkar, Nicholas Moore, Nicole Memblatt, Nikki Miocevic,

Nirav Shah, Nitin Kumar, Olga Herrera, Patrick Floody, Paul D'Ambrosio, Qin Li, Rafia Bhore, Rakiyya Ali, Ramya Iyer, Ravikanth Chava, Rinol Alaj, Romana Hosain, Ruchin Gorawala, Ryan Yu, Rylee Fogarty, S. Balachandra Dass, Sagarika Bollini, Samit Ganguly, Sandra DeCicco, Sara Dale, Sara Hamon, Sarah Cassimaty, Selin Somersan-Karakaya, Shane McCarthy, Sharon Henkel, Shazia Ali, Soraya Nossoughi, Steven Elkin, Sumathi Sivapalasingam, Susan Irvin, Tami Min, Ted Burczynski, Theresa Devins, Thomas Norton, Travis Bernardo, Vinh Nuce, Wilson Caldwell, Yanmei Tian, Yasmin Khan

### Supplementary Methods

#### Additional Statistical Methods

##### Virologic Endpoint

The virologic efficacy endpoint of time-weighted average (TWA) daily change from baseline (day 1) through day 7 was calculated for each patient as the area under the viral load–time curve with the use of the linear trapezoidal rule (area under the curve for change from baseline divided by the time interval [in days] of the observation period) and analyzed using an analysis of covariance model with treatment group, country, and risk factor (no risk factor vs at least one risk factor) as fixed effects and baseline viral load and treatment by baseline interaction as covariates.

##### Clinical Endpoints

The proportions of patients with medically attended visits (MAVs) due to worsening COVID-19 were compared between the REGEN-COV combined dose group and placebo as well as between each REGEN-COV treatment arm and placebo using Fisher's exact test at a two-sided alpha level of 0.05. A similar analysis was performed for the proportion of patients with COVID-19–related hospitalization or emergency room or urgent care visits, as well as for the proportions of patients with each type of MAV.

##### Missing Data Handling

Missing data for virology endpoints were handled as follows: Analysis-positive polymerase chain reaction (PCR) results below the lower limit of quantification (LLOQ) of 714 copies/mL (2.85 log10 copies/mL) were imputed as half the LLOQ (357 copies/mL), and negative PCR results were imputed as 0 $log_{10}$ copies/mL (1 copy/mL). PCR results greater than the upper limit of quantification of $7.1x10^7$ copies/mL (7.85 $log_{10}$ copies/mL) were imputed as $7.1x10^7$ copies/mL (7.85 $log_{10}$ copies/mL).

For categorical variables, patients with missing data were included in calculations of percentages.

#### Measurement of REGN10933 and REGN10987 in Serum

Serum for drug concentration analysis was collected from all patients at pre-dose (at the screening or baseline visit), day 1 at the end of the infusion, and day 29. Additional serum collections were on days 3, 5, 7, and 15 for phase 1 patients only.

The human serum concentrations of REGN10933 (casirivimab) and REGN10987 (imdevimab) were measured using validated immunoassays which employ streptavidin microplates from Meso Scale Discovery (MSD, Gaithersburg, MD, USA). The methods utilized two anti-idiotypic monoclonal antibodies, each specific for either REGN10933 or REGN10987, as the capture antibodies. Captured REGN10933 and REGN10987 were detected using two different, non-competing anti-idiotypic monoclonal antibodies, each also specific for either REGN10933 or REGN10987. The bioanalytical methods specifically quantitated the levels of each anti–SARS-CoV-2 spike monoclonal antibody separately, with no interference from the other antibody. The assay has an LLOQ of 0.156 µg/mL for each analyte in the undiluted serum sample.

#### Supplementary Figures

##### Figure S1. Schematic Overview of the Study Design

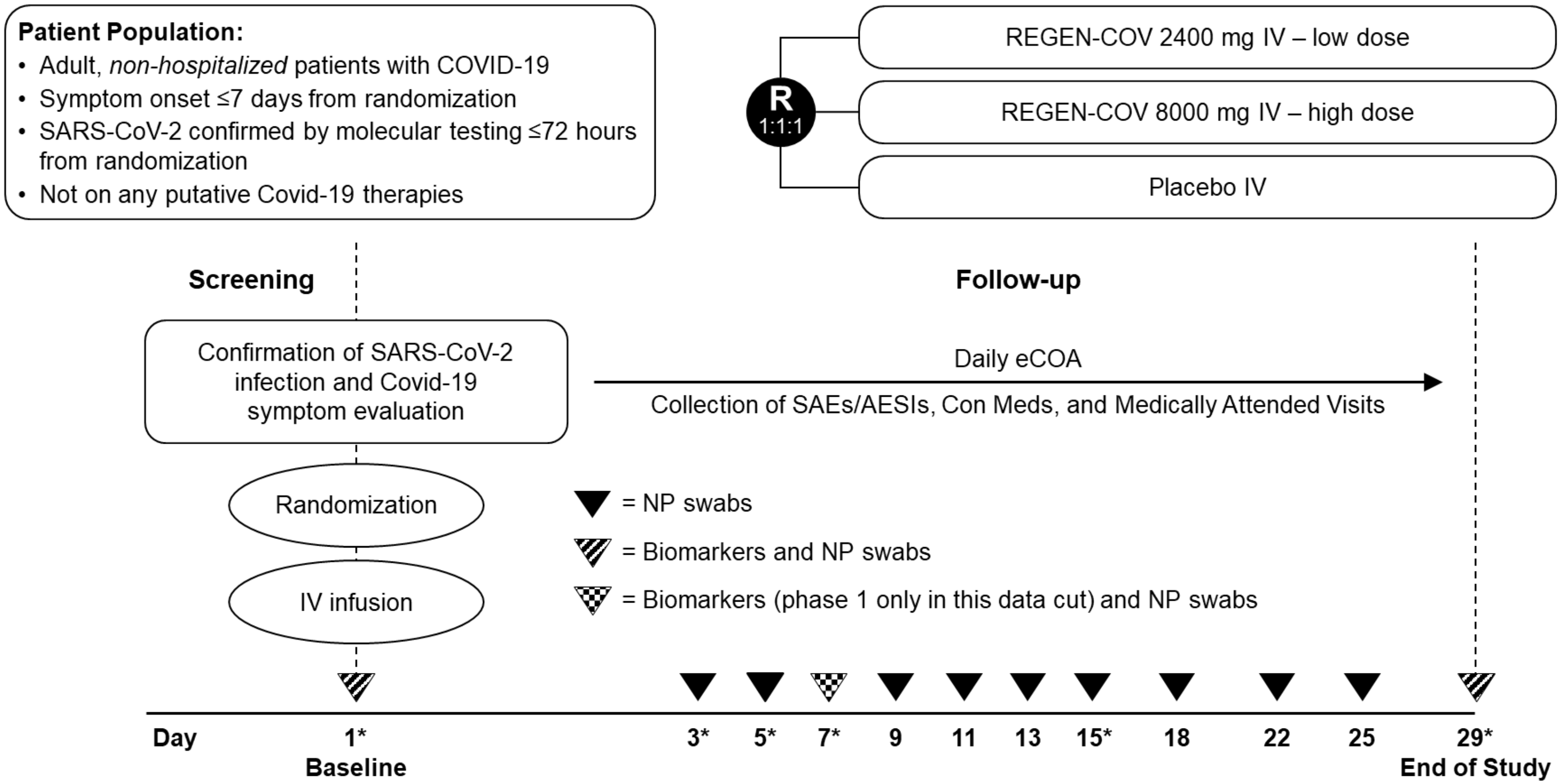

* Serum samples for pharmacokinetic analysis were collected on day 3, 5, 7, and 15 in the phase 1 part only.

Abbreviations: AESI, adverse event of special interest; con med, concomitant medication; eCOA, electronic clinical outcome assessment; IV, intravenous(ly); NP, nasopharyngeal; R, randomized; SAE, serious adverse event.

#### Figure S2. Relationship Between Baseline Serum Antibody Status and Baseline Viral Load

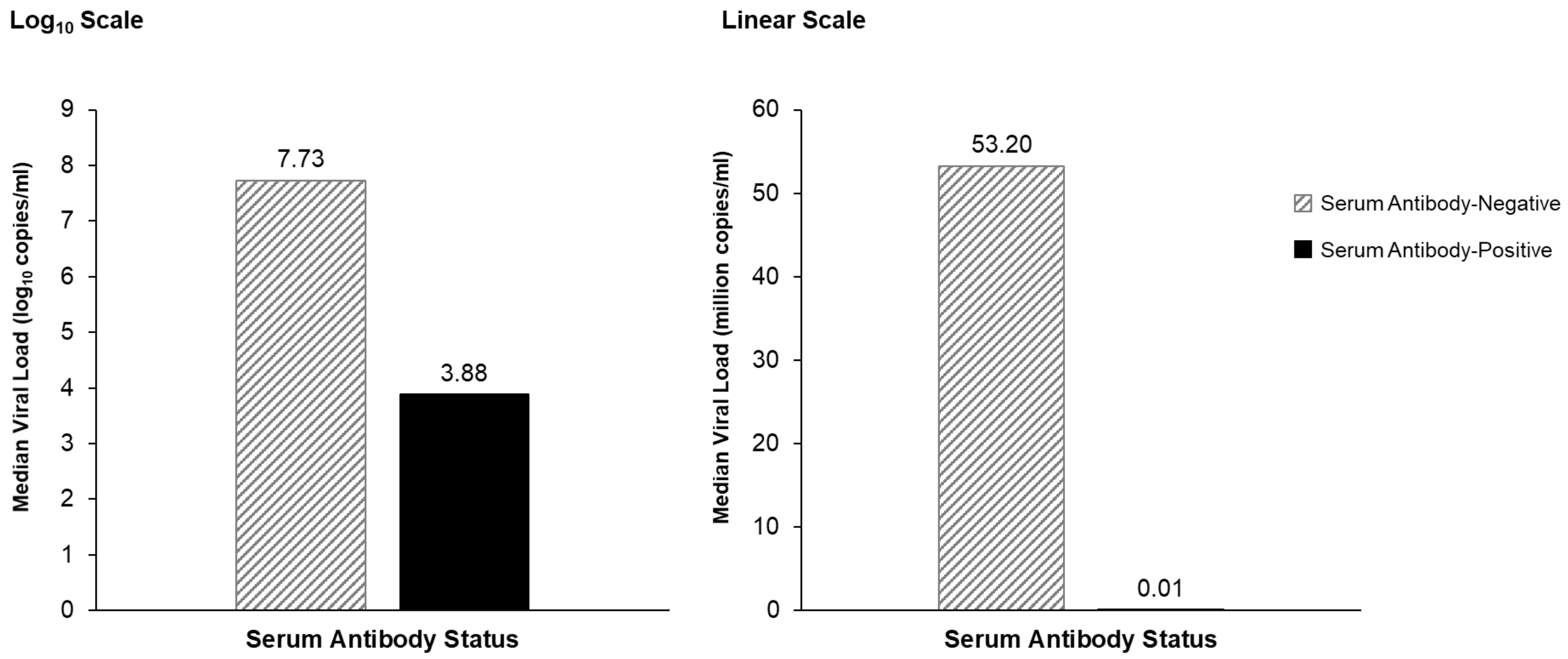

| **No. of Patients by Serum Antibody Status** | **Median (Q1, Q3) Viral Load in Nasoparyngeal Swab, $Log_{10}$ Scale by Serum Antibody Status:** | **Median (Q1, Q3) Viral Load in Nasoparyngeal Swab, Linear Scale by Serum Antibody Status:** | **Median Time of Covid-19 Symptoms Before Randomization**: 3.0 days |
|---|---|---|---|
| Negative: 292/524 (55.7%) | Negative: 7.73 (6.28, 7.85) $log_{10}$ copies/ml | Negative: 53.20 (1.91, 71.00) million copies/ml | |
| Positive: 176/524 (33.6%) | Positive: 3.88 (3.18, 5.55) $log_{10}$ copies/ml | Positive: 0.01 (0.00, 0.35) million copies/ml | |
| Other: 56/524 (10.7%) | | | |

Abbreviation: Q, quartile.

**Figure S3. Viral Load Over Time in the Placebo Arm by Baseline Serum Antibody Status**

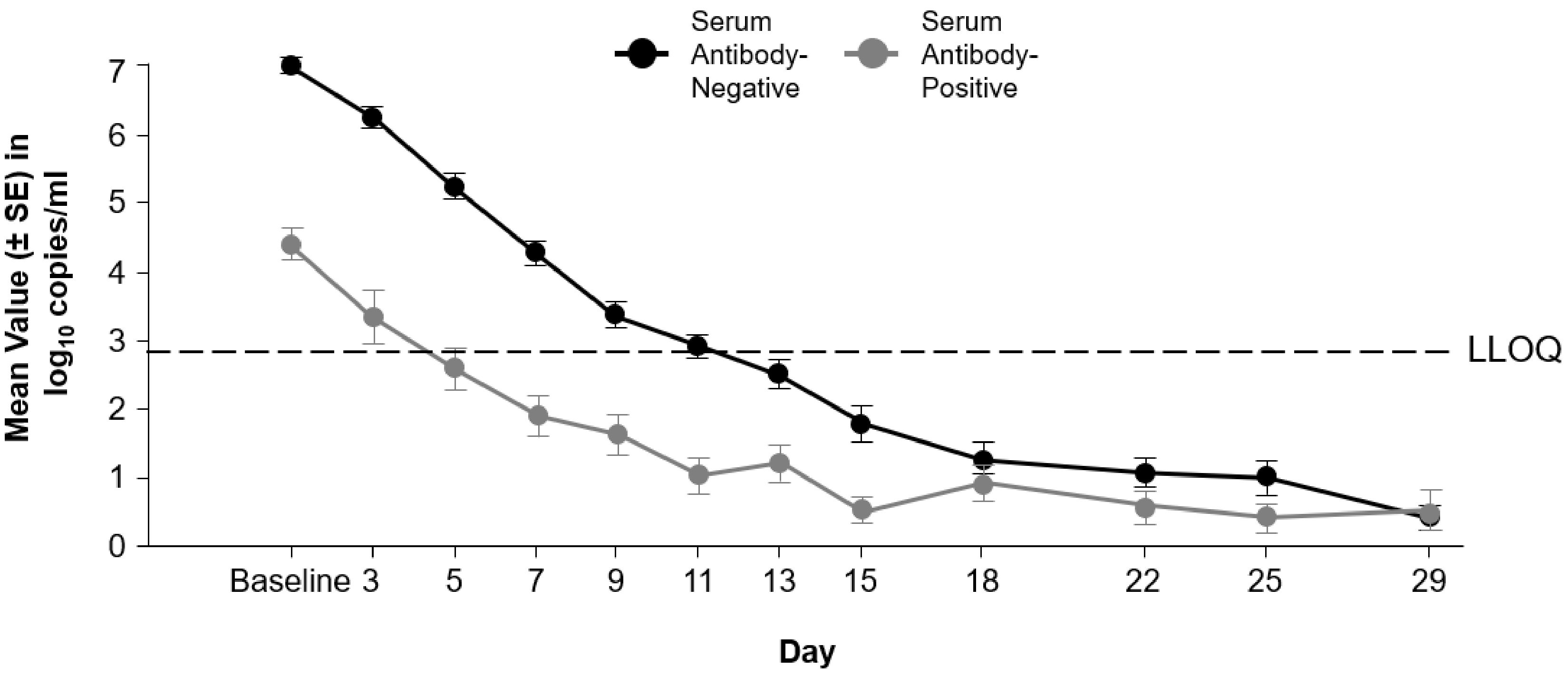

Abbreviations: LLOQ, lower limit of quantification; SE, standard error.

**Figure S4. Proportion of Patients in the Placebo Arm With ≥1 COVID-19–Related Medically Attended Visit Through Day 29**

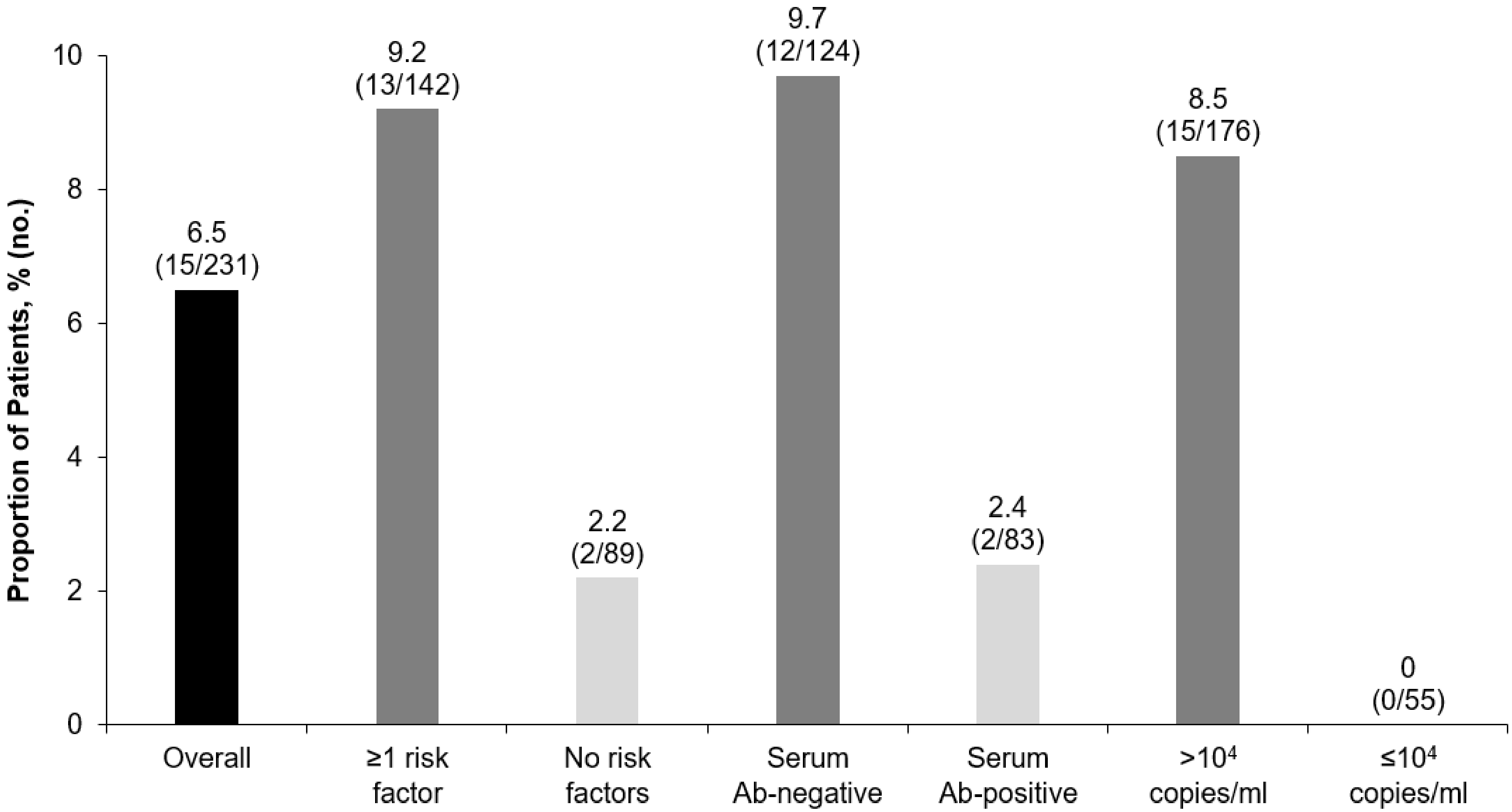

Abbreviation: Ab, antibody.

#### Figure S5. Time-weighted Average Daily Change from Baseline in Viral Load ($log_{10}$ copies/ml) with REGEN-COV Treatment – Forest Plots

##### A. Average Daily Viral Load ($log_{10}$ copies/mL) Through Day 7 (Analysis Group 2)

| Group | REGEN-COV | Placebo | Difference (95% CI) | P Value |
|---|---|---|---|---|
| | Time-weighted average change Mean (Standard Error) | | | |
| **Baseline Viral Load >10^7 copies/mL** | | | | |
| Combined dose groups vs Placebo (n=185) | -2.13 (0.13) | -1.46 (0.15) | -0.68 (-0.94, -0.41) | <0.0001 |
| 8000 mg vs Placebo (n=124) | -2.13 (0.16) | -1.46 (0.15) | -0.68 (-0.99, -0.36) | <0.0001 |
| 2400 mg vs Placebo (n=132) | -2.14 (0.16) | -1.46 (0.15) | -0.68 (-0.99, -0.37) | <0.0001 |
| **Baseline Viral Load >10^6 copies/mL** | | | | |
| Combined dose groups vs Placebo (n=237) | -2.06 (0.11) | -1.40 (0.13) | -0.65 (-0.89, -0.41) | <0.0001 |
| 8000 mg vs Placebo (n=161) | -1.98 (0.13) | -1.40 (0.13) | -0.58 (-0.86, -0.30) | <0.0001 |
| 2400 mg vs Placebo (n=163) | -2.13 (0.13) | -1.40 (0.13) | -0.73 (-1.01, -0.45) | <0.0001 |
| **Baseline Seronegative** | | | | |
| Combined dose groups vs Placebo (n=256) | -1.91 (0.08) | -1.18 (0.10) | -0.73 (-0.97, -0.48) | <0.0001 |
| **Baseline Seropositive** | | | | |
| Combined dose groups vs Placebo (n=137) | -1.32 (0.10) | -1.37 (0.16) | 0.05 (-0.32, 0.42) | 0.7866 |
| **mFAS** | | | | |
| **Combined dose groups vs Placebo (n=437)** | **-1.66 (0.07)** | **-1.30 (0.09)** | **-0.36 (-0.56, -0.16)** | **0.0003** |

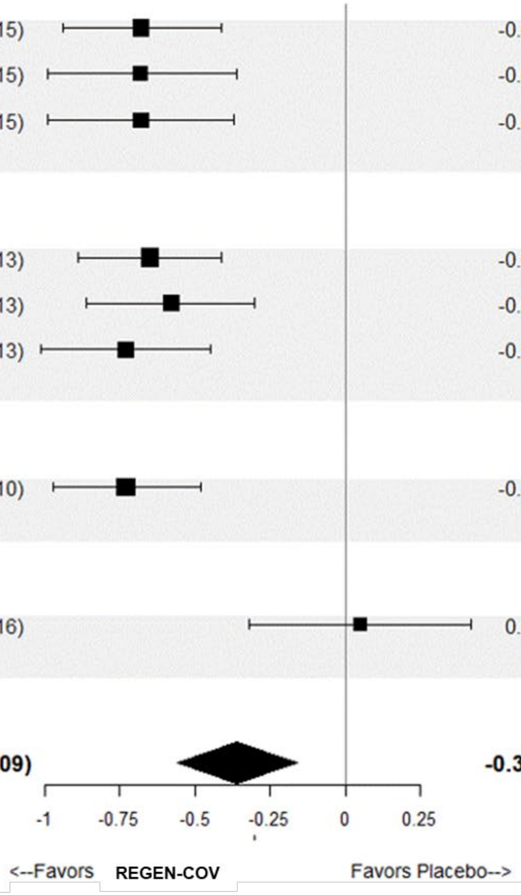

**B. Average Daily Viral Load ($\log_{10}$ copies/mL) Through Day 7 (Analysis Groups 1+2)**

| Group | REGEN-COV | Placebo | Difference (95% CI) | P Value |
|---|---|---|---|---|
| | Time-weighted average change Mean (Standard Error) | | | |
| **Baseline Viral Load >10^7 copies/mL** | | | | |
| Combined dose groups vs Placebo (n=256) | -3.16 (0.12) | -2.37 (0.14) | -0.80 (-1.05, -0.55) | <0.0001 |
| 8000 mg vs Placebo (n=175) | -3.22 (0.14) | -2.37 (0.14) | -0.85 (-1.14, -0.56) | <0.0001 |
| 2400 mg vs Placebo (n=174) | -3.13 (0.14) | -2.37 (0.14) | -0.76 (-1.05, -0.47) | <0.0001 |
| **Baseline Viral Load >10^6 copies/mL** | | | | |
| Combined dose groups vs Placebo (n=332) | -3.04 (0.10) | -2.29 (0.12) | -0.75 (-0.98, -0.52) | <0.0001 |
| 8000 mg vs Placebo (n=222) | -3.03 (0.12) | -2.29 (0.12) | -0.75 (-1.01, -0.48) | <0.0001 |
| 2400 mg vs Placebo (n=224) | -3.04 (0.12) | -2.29 (0.12) | -0.75 (-1.02, -0.49) | <0.0001 |
| **Baseline Seronegative** | | | | |
| Combined dose groups vs Placebo (n=360) | -2.72 (0.07) | -2.03 (0.10) | -0.69 (-0.91, -0.46) | <0.0001 |
| **Baseline Seropositive** | | | | |
| Combined dose groups vs Placebo (n=236) | -1.83 (0.08) | -1.92 (0.11) | 0.09 (-0.18, 0.36) | 0.5215 |
| **mFAS** | | | | |
| **Combined dose groups vs Placebo (n=665)** | **-2.36 (0.06)** | **-2.01 (0.08)** | **-0.34 (-0.51, -0.18)** | **<0.0001** |

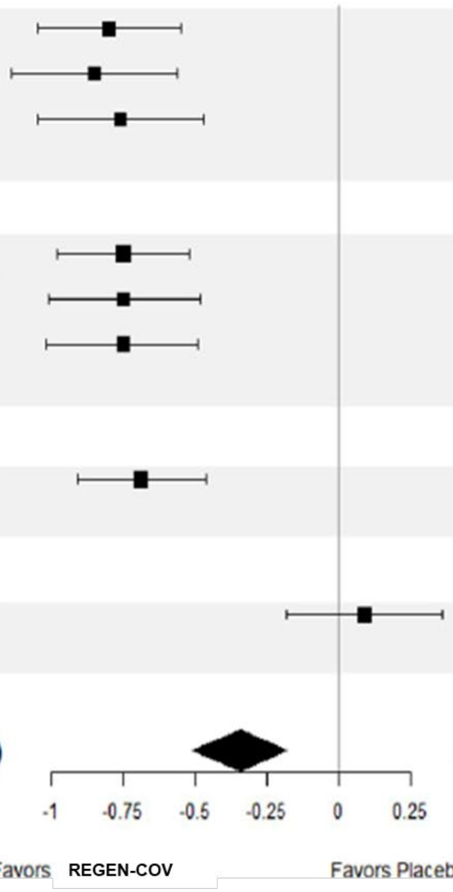

Adjusted analysis of time-weighted average daily change from baseline in viral load ($\log_{10}$ copies/mL) was based on analysis of covariance (ANCOVA) model with treatment group, country, risk factor, and baseline serology status as fixed effects and on baseline viral load and treatment by baseline viral load as covariates.
Abbreviations: CI, confidence interval; mFAS, modified full analysis set.

### Figure S6. Time-Weighted Average Daily Change from Baseline in Viral Load ($log_{10}$ copies/mL) With REGEN-COV Treatment – Graphs

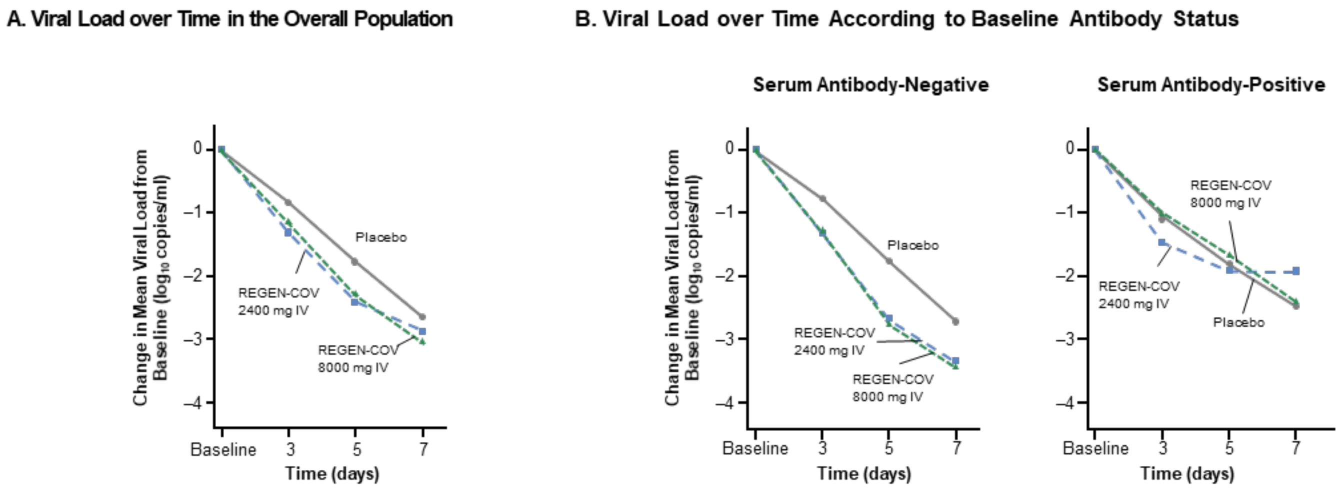

**No. at Risk**

| | | | | |
|---|---|---|---|---|
| Placebo | 150 | 136 | 143 | 146 |
| REGEN-COV 2400 mg IV | 142 | 132 | 137 | 137 |
| REGEN-COV 8000 mg IV | 145 | 135 | 140 | 141 |

**No. at Risk**

| | | | | | | | | |
|---|---|---|---|---|---|---|---|---|
| Placebo | 94 | 87 | 91 | 93 | 42 | 39 | 40 | 41 |
| REGEN-COV 2400 mg IV | 83 | 78 | 80 | 80 | 45 | 42 | 45 | 45 |
| REGEN-COV 8000 mg IV | 79 | 73 | 76 | 77 | 50 | 49 | 50 | 50 |

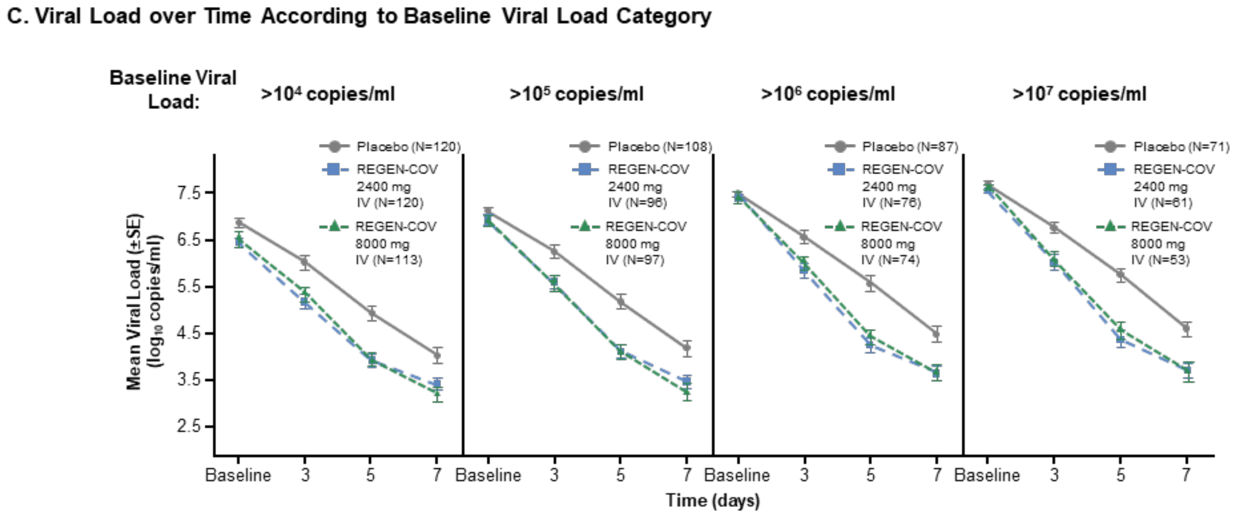

Shown is the change in mean viral load (in $log_{10}$ copies/mL) from baseline at each visit through day 7 in the overall population (mFAS, which excluded patients who tested negative for severe acute respiratory syndrome coronavirus 2 by qualitative reverse-transcriptase polymerase chain reaction at baseline) and in groups defined according to baseline antibody status and baseline viral load. I bars in Panel C indicate the standard error. The lower limit of detection (dashed line) is 714 copies/ml (2.85 $log_{10}$ copies/mL).

Abbreviations: IV, intravenous(ly); mFAS, modified full analysis set; SE, standard error.

### Figure S7. Time to Sustained Negative RT-qPCR by Baseline Viral Load Category

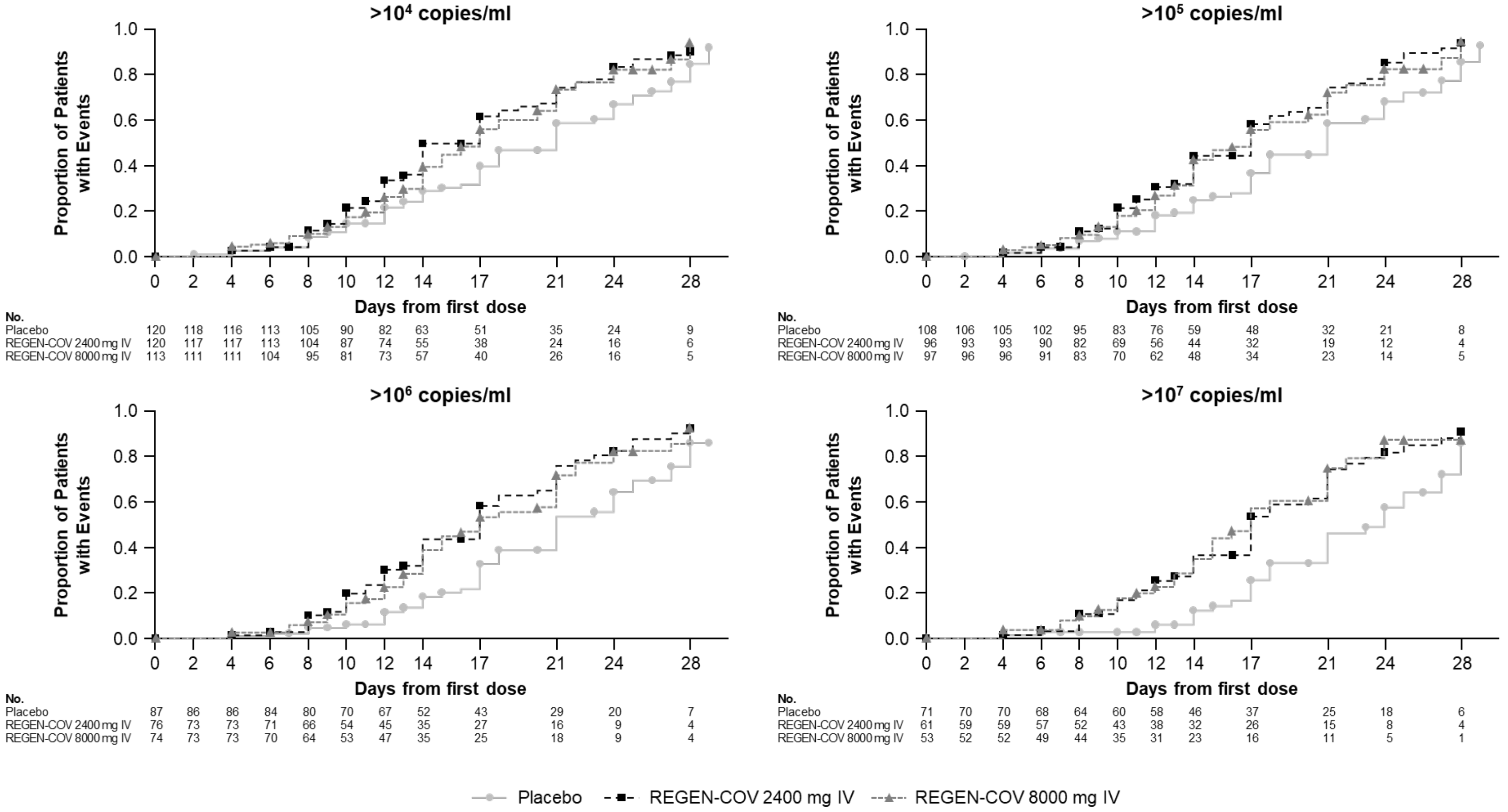

The time to negative RT-qPCR in NP swabs with no subsequent positive RT-qPCR for analysis group 2 is shown above.

Abbreviations: IV, intravenous(ly); NP, nasopharyngeal; RT-qPCR, quantitative reverse-transcriptase polymerase chain reaction.

### Figure S8. Proportion of Patients With High Viral Load at Each Visit

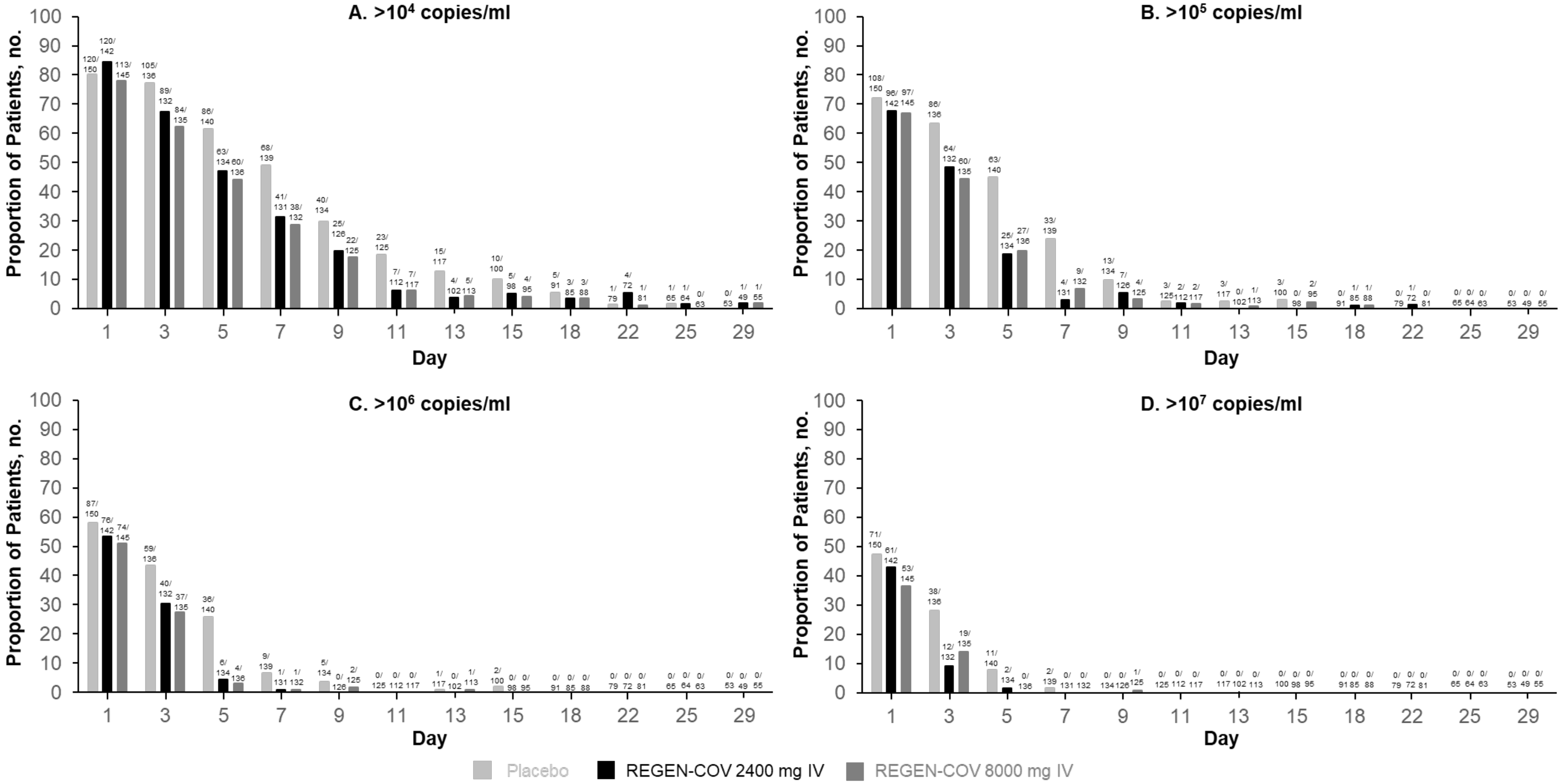

The proportion of patients at each visit with a viral load greater than $10^4$ (A) $10^5$ (B) $10^6$ (C) $10^7$ (D) is shown above.

Abbreviation: IV, intravenous(ly).

#### Figure S9. Viral Load Through Day 29 in Patients With and Without ≥1 COVID-19–Related Medically Attended Visit

##### A. Viral Load Over Time in Patients With and Without ≥1 COVID-19–Related MAV

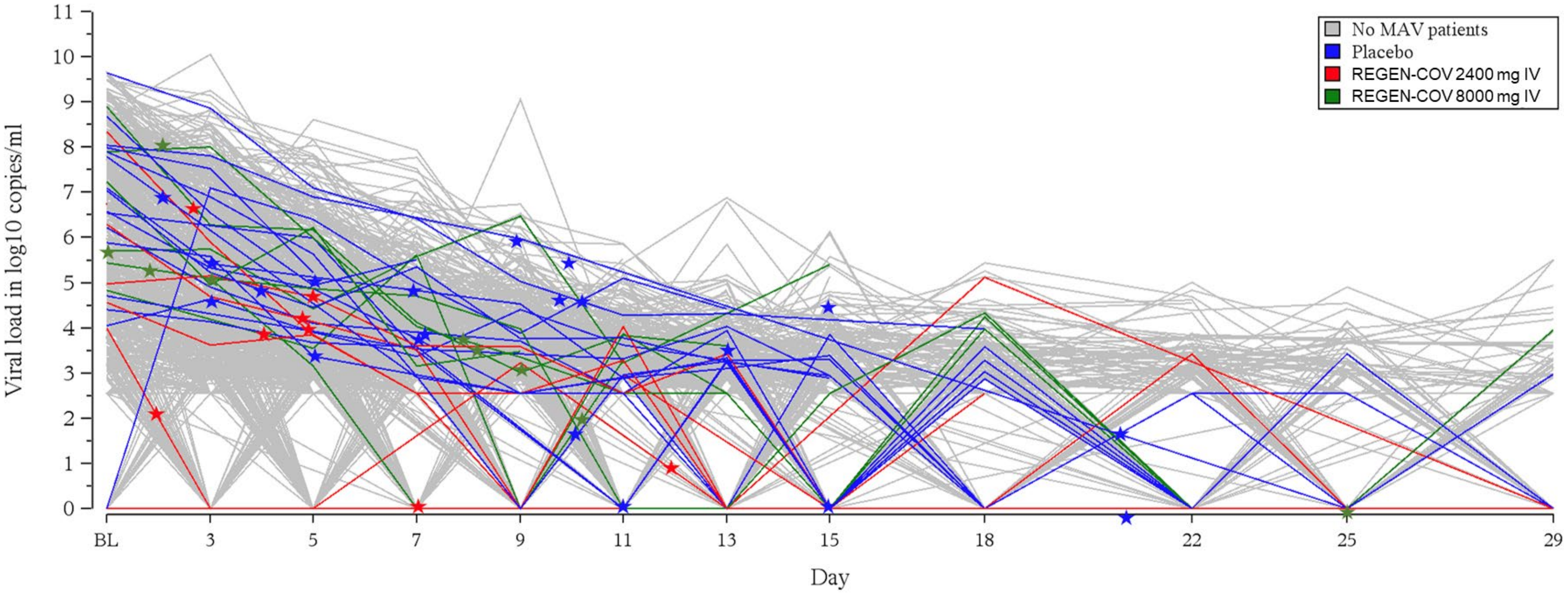

Abbreviations: IV, intravenously; MAV, medically attended visit.

**B. Viral Load Over Time in Patients With ≥1 COVID-19–Related MAV – Placebo**

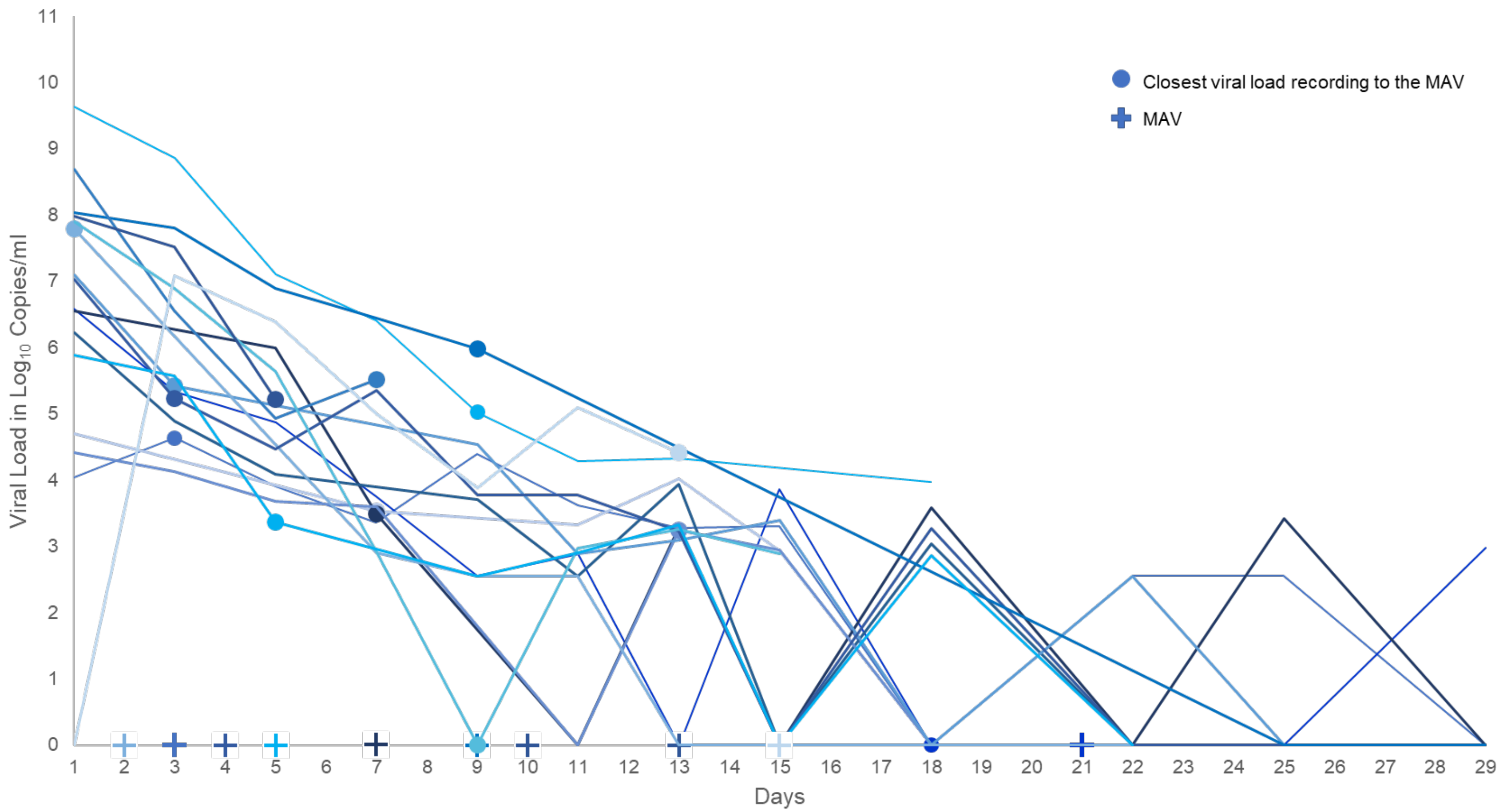

Abbreviation: MAV, medically attended visit.

**C. Viral Load Over Time in Patients With ≥1 COVID-19–Related MAV – REGEN-COV**

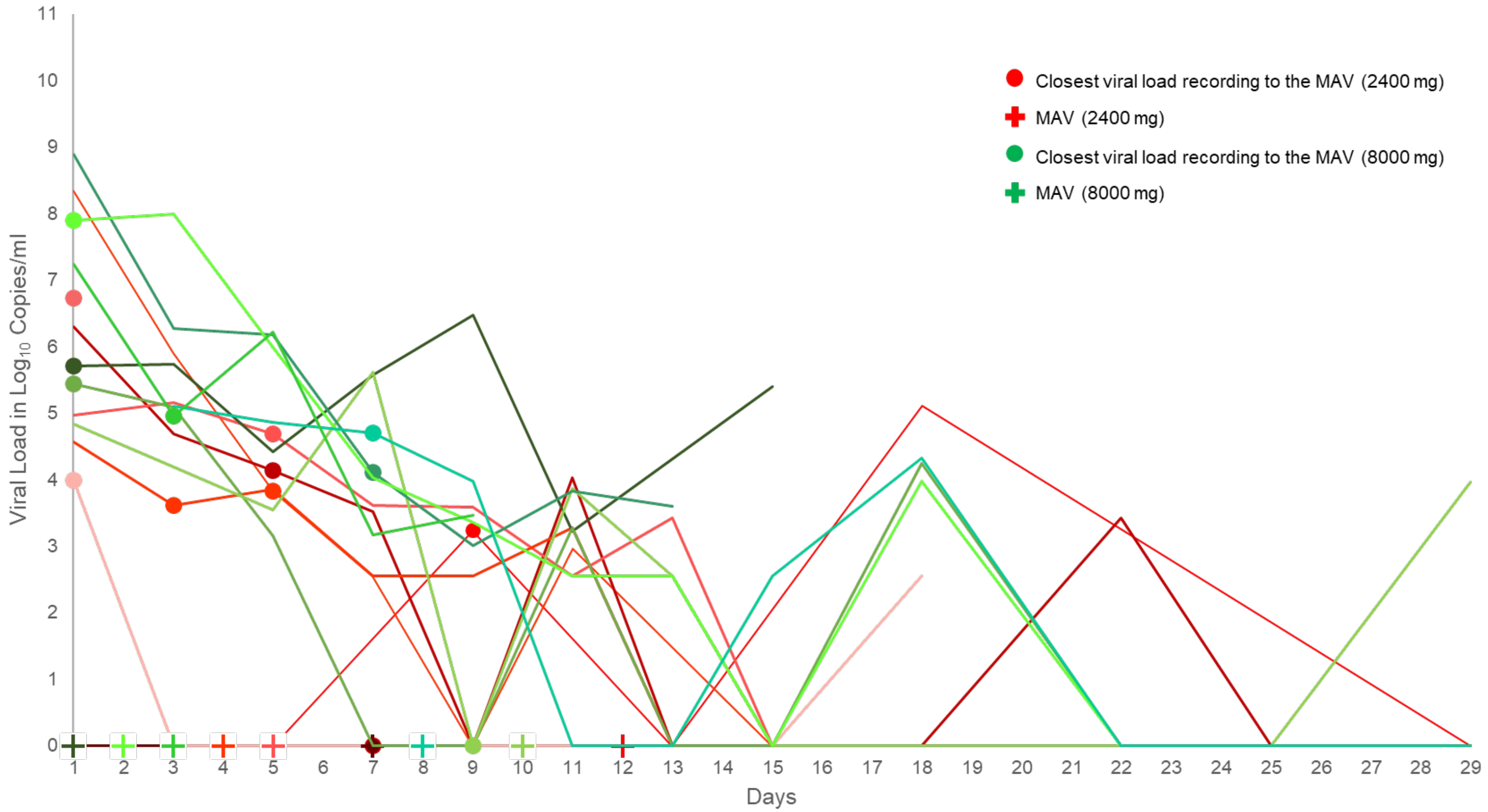

Abbreviation: MAV, medically attended visit.

### Supplementary Tables

#### Table S1. Phase 1/2 Primary Analysis of Virologic and Clinical Endpoints

The primary analysis of virologic endpoints and clinical endpoints was conducted at a two-sided α = 0.05. Eight virologic endpoints and two clinical endpoints were tested hierarchically in the following order:

| Endpoint Number | Description |
| --- | --- |
| 1 | Time-weighted average daily change from baseline in viral load ($log_{10}$ copies/mL) from day 1 through day 7 in the mFAS patients with baseline viral load >$10^7$ copies/mL for REGEN-COV 2400 mg and 8000 mg combined group versus placebo (patients 276 through 799) |
| 2 | Time-weighted average daily change from baseline in viral load ($log_{10}$ copies/ml) from day 1 through day 7 in the mFAS patients with baseline viral load >$10^6$ copies/mL for REGEN-COV 2400 mg and 8000 mg combined group versus placebo (patients 276 through 799) |
| 3 | Time-weighted average daily change from baseline in viral load ($log_{10}$ copies/mL) from day 1 through day 7 in the serum antibody–negative (seronegative) mFAS for REGEN-COV 2400 mg and 8000 mg combined group versus placebo (patients 276 through 799) |
| 4 | Time-weighted average daily change from baseline in viral load ($log_{10}$ copies/ml) from day 1 through day 7 in the mFAS for REGEN-COV 2400 mg and 8000 mg combined group versus placebo (patients 276 through 799) |
| 5 | Time-weighted average daily change from baseline in viral load ($log_{10}$ copies/mL) from day 1 through day 7 in the mFAS patients with baseline viral load >$10^7$ copies/mL for REGEN-COV 8000 mg group versus placebo (patients 276 through 799) |
| 6 | Time-weighted average daily change from baseline in viral load ($log_{10}$ copies/mL) from day 1 through day 7 in the mFAS patients with baseline viral load >$10^7$ copies/mL for REGEN-COV 2400 mg group versus placebo (patients 276 through 799) |
| 7 | Time-weighted average daily change from baseline in viral load ($log_{10}$ copies/mL) from day 1 through day 7 in the mFAS patients with baseline viral load >$10^6$ copies/mL for REGEN-COV 8000 mg group versus placebo (patients 276 through 799) |
| 8 | Time-weighted average daily change from baseline in viral load ($log_{10}$ copies/mL) from day 1 through day 7 in the mFAS patients with baseline viral load >$10^6$ copies/mL for REGEN-COV 2400 mg versus placebo (patients 276 through 799) |

| Endpoint Number | Description |
|---|---|
| 9 | Proportion of patients with MAVs through day 29 in the mFAS for REGEN-COV 2400 mg and 8000 mg combined group versus placebo (patients 1 through 799) |
| 10 | Proportion of patients with subset of MAVs consisting only of hospitalization or emergency room visit or urgent care visit through day 29 in the mFAS for REGEN-COV 2400 mg and 8000 mg combined group versus placebo (patients 1 through 799) |

Abbreviations: mFAS, modified full analysis set; MAV, medically attended visit.

#### Table S2. Demographic and Baseline Medical Characteristics (Group-2; Full Analysis Set)

**Full analysis set (group-2): Patients who underwent randomization in Phase 2**

| Characteristic | Total (n = 524) | Placebo (n = 173) | REGEN-COV 2400 mg (n = 174) | REGEN-COV 8000 mg (n = 177) | REGEN-COV combined (n = 351) |
|---|---|---|---|---|---|
| Age, y, median (IQR) | 41.0 (28.0–52.0) | 40.0 (28.0–53.0) | 42.0 (29.0–53.0) | 39.0 (28.0–51.0) | 41.0 (28.0–52.0) |
| Male sex | 242 (46.2) | 84 (48.6) | 76 (43.7) | 82 (46.3) | 158 (45.0) |
| Hispanic or Latino ethnic group[a] | 250 (47.7) | 91 (52.6) | 80 (46.0) | 79 (44.6) | 159 (45.3) |
| Race[a] | | | | | |
| White | 457 (87.2) | 155 (89.6) | 150 (86.2) | 152 (85.9) | 302 (86.0) |
| Black or African American | 39 (7.4) | 10 (5.8) | 12 (6.9) | 17 (9.6) | 29 (8.3) |
| Asian | 11 (2.1) | 2 (1.2) | 6 (3.4) | 3 (1.7) | 9 (2.6) |
| Native American or Alaska Native | 3 (0.6) | 1 (0.6) | 1 (0.6) | 1 (0.6) | 2 (0.6) |
| Unknown | 4 (0.8) | 1 (0.6) | 1 (0.6) | 2 (1.1) | 3 (0.9) |
| Not reported | 10 (1.9) | 4 (2.3) | 4 (2.3) | 2 (1.1) | 6 (1.7) |
| Weight, kg, median (IQR) | 79.80 (68.00–93.00) | 81.00 (69.40–91.00) | 79.30 (68.35–91.40) | 79.40 (67.60–95.70) | 79.40 (68.00–94.00) |

| | | | | | |
|---|---|---|---|---|---|
| Body-mass index (SD)[b] | 29.38 (8.800) | 30.07 (11.828) | 29.03 (6.291) | 29.04 (7.386) | 29.04 (6.855) |
| Obesity[c] | 183 (34.9) | 59 (34.1) | 62 (35.6) | 62 (35.0) | 124 (35.3) |
| Baseline viral load in nasopharyngeal swab (raw values) | | | | | |
| No. of patients | 494 | 168 | 159 | 167 | 326 |
| Viral load, mean, copies/mL (SD) | 20153955.3 (28853829.01) | 25281106.2 (31606803.98) | 18101836.4 (26953942.48) | 16949916.6 (27112316.53) | 17511742.5 (26999735.08) |
| Viral load, median, copies/mL (range) | 630500.0 (1–71000000) | 1660000.0 (1–71000000) | 301000.0 (1–71000000) | 369000.0 (1–71000000) | 342500.0 (1–71000000) |
| Baseline viral load in nasopharyngeal swab ($log_{10}$ scale) | | | | | |
| No. of patients | 494 | 168 | 159 | 167 | 326 |
| Viral load, mean, $log_{10}$ copies/mL (SD) | 5.30 (2.514) | 5.50 (2.546) | 5.34 (2.426) | 5.08 (2.560) | 5.20 (2.495) |
| Viral load, median, $log_{10}$ copies/mL (range) | 5.80 (0.0–7.9) | 6.22 (0.0–7.9) | 5.48 (0.0–7.9) | 5.57 (0.0–7.9) | 5.53 (0.0–7.9) |
| Baseline viral load in nasopharyngeal swab category | | | | | |
| $>10^4$ | 353 (67.4) | 120 (69.4) | 120 (69.0) | 113 (63.8) | 233 (66.4) |
| $>10^5$ | 301 (57.4) | 108 (62.4) | 96 (55.2) | 97 (54.8) | 193 (55.0) |

| | | | | | |
|---|---|---|---|---|---|
| >$10^6$ | 237 (45.2) | 87 (50.3) | 76 (43.7) | 74 (41.8) | 150 (42.7) |
| >$10^7$ | 185 (35.3) | 71 (41.0) | 61 (35.1) | 53 (29.9) | 114 (32.5) |
| Positive baseline qualitative RT-PCR | 437 (83.4) | 150 (86.7) | 142 (81.6) | 145 (81.9) | 287 (81.8) |
| Baseline serum C-reactive protein level | | | | | |
| No. of patients | 333 | 111 | 112 | 110 | 222 |
| Level, mean, mg/L (SD) | 13.066 (25.5848) | 17.722 (31.9234) | 10.931 (20.8046) | 10.540 (22.1667) | 10.738 (21.4425) |
| Level, median, mg/L (range) | 3.570 (0.10–157.96) | 5.020 (0.10–153.80) | 3.640 (0.12–135.36) | 2.475 (0.18–157.96) | 3.145 (0.12–157.96) |
| Baseline serum antibody status | | | | | |
| Negative | 292 (55.7) | 101 (58.4) | 96 (55.2) | 95 (53.7) | 191 (54.4) |
| Positive | 176 (33.6) | 56 (32.4) | 58 (33.3) | 62 (35.0) | 120 (34.2) |
| Other | 56 (10.7) | 16 (9.2) | 20 (11.5) | 20 (11.3) | 40 (11.4) |
| Time from symptom onset to randomization, median, d (IQR) | 3.0 (2.0–5.0) | 3.0 (2.0–5.0) | 3.0 (2.0–5.0) | 3.0 (2.0–5.0) | 3.0 (2.0–5.0) |
| At least one risk factor for hospitalization[d] | 307 (58.6) | 100 (57.8) | 108 (62.1) | 99 (55.9) | 207 (59.0) |

Data are presented as no. (%) unless otherwise indicated. Percentages may not total 100 because of rounding.
[a]Race and ethnic group were self-reported by the patients.
[b]The body-mass index is the weight in kilograms divided by the square of the height in meters.
[c]Obesity is defined as a body-mass index of greater than 30.

[d]Risk factors for hospitalization include an age of more than 50 years, obesity, cardiovascular disease (including hypertension), chronic lung disease (including asthma), chronic metabolic disease (including diabetes), chronic kidney disease (including receipt of dialysis), chronic liver disease, and immunocompromised (immunosuppression or receipt of immunosuppressants).
Abbreviations: IQR, interquartile range; RT-PCR, reverse-transcriptase polymerase chain reaction; SD, standard deviation.

#### Table S3. Demographic and Baseline Medical Characteristics (Group-1+2; Modified Full Analysis Set)

**Modified full analysis set (group-1+2): Patients with detectable SARS-CoV-2 in baseline nasopharyngeal sample by central lab RT-qPCR who underwent randomization in Phase 1 and 2**

| Characteristic | Total (n = 665) | Placebo (n = 231) | REGEN-COV 2400 mg (n = 215) | REGEN-COV 8000 mg (n = 219) | REGEN-COV combined (n = 434) |
|---|---|---|---|---|---|
| Age, y, median (IQR) | 42.0 (31.0–52.0) | 41.0 (32.0–53.0) | 42.0 (30.0–52.0) | 42.0 (30.0–52.0) | 42.0 (30.0–52.0) |
| Male sex | 316(47.5) | 121 (52.4) | 94 (43.7) | 101 (46.1) | 195 (44.9) |
| Hispanic or Latino ethnic group[a] | 321 (48.3) | 114 (49.4) | 98 (45.6) | 109 (49.8) | 207 (47.7) |
| Race[a] | | | | | |
| White | 556 (85.1) | 198 (85.7) | 181 (84.2) | 187 (85.4) | 368 (84.8) |
| Black or African American | 64 (9.6) | 20 (8.7) | 23 (10.7) | 21 (9.6) | 44 (10.1) |
| Asian | 10 (1.5) | 3 (1.3) | 5 (2.3) | 2 (0.9) | 7 (1.6) |
| Native American or Alaska Native | 4 (0.6) | 2 (0.9) | 1 (0.5) | 1 (0.5) | 2 (0.5) |
| Unknown | 5 (0.8) | 3 (1.3) | 0 | 2 (0.9) | 2 (0.5) |
| Not reported | 16 (2.4) | 5 (2.2) | 5 (2.3) | 6 (2.7) | 11 (2.5) |
| Weight, kg, median (IQR) | 81.60 (69.40–95.30) | 83.00 (71.00–94.90) | 81.20 (69.30–94.20) | 81.60 (68.80–97.00) | 81.20 (68.90–95.30) |

| | | | | | |
|---|---|---|---|---|---|
| Body-mass index (SD)[b] | 29.68 (8.545) | 30.13 (10.950) | 29.37 (6.593) | 29.53 (7.354) | 29.45 (6.979) |
| Obesity[c] | 252 (37.9) | 84 (36.4) | 80 (37.2) | 88 (40.2) | 168 (38.7) |
| Baseline viral load in nasopharyngeal swab (raw values) | | | | | |
| No. of patients | 665 | 231 | 215 | 219 | 434 |
| Viral load, mean, copies/mL (SD) | 21168074.1 (29385726.53) | 23489191.6 (31004393.79) | 19670523.0 (28151618.29) | 20189970.5 (28794678.24) | 19932640.5 (28446225.30) |
| Viral load, median, copies/mL (range) | 984000.0 (357–71000000) | 898000.0 (357–71000000) | 1450000.0 (357–71000000) | 868000.0 (357–71000000) | 1130000.0 (357–71000000) |
| Baseline viral load in nasopharyngeal swab ($log_{10}$ scale) | | | | | |
| No. of patients | 665 | 231 | 215 | 219 | 434 |
| Viral load, mean, $log_{10}$ copies/mL (SD) | 5.84 (1.766) | 5.84 (1.835) | 5.92 (1.653) | 5.77 (1.804) | 5.84 (1.730) |
| Viral load, median, $log_{10}$ copies/mL (range) | 5.99 (2.6–7.9) | 5.95 (2.6–7.9) | 6.16 (2.6–7.9) | 5.94 (2.6–7.9) | 6.05 (2.6–7.9) |
| Baseline viral load in nasopharyngeal swab category | | | | | |
| $>10^4$ | 523 (78.6) | 176 (76.2) | 180 (83.7) | 167 (76.3) | 347 (80.0) |
| $>10^5$ | 439 (66.0) | 149 (64.5) | 148 (68.8) | 142 (64.8) | 290 (66.8) |

| | | | | | |
|---|---|---|---|---|---|
| $>10^6$ | 332 (49.9) | 114 (49.4) | 110 (51.2) | 108 (49.3) | 218 (50.2) |
| $>10^7$ | 256 (38.5) | 93 (40.3) | 82 (38.1) | 81 (37.0) | 163 (37.6) |
| Positive baseline qualitative RT-PCR | 665 (100) | 231 (100) | 215 (100) | 219 (100) | 434 (100) |
| Baseline serum C-reactive protein level | | | | | |
| No. of patients | 503 | 178 | 161 | 164 | 325 |
| Level, mean, mg/L (SD) | 14.667 (29.7365) | 19.886 (37.1774) | 11.631 (25.9227) | 11.983 (22.7314) | 11.809 (24.3275) |
| Level, median, mg/L (range) | 4.000 (0.10–239.67) | 4.900 (0.10–232.04) | 3.400 (0.12–239.67) | 3.575 (0.14–157.96) | 3.570 (0.12–239.67) |
| Baseline serum antibody status | | | | | |
| Negative | 360 (54.1) | 124 (53.7) | 121 (56.3) | 115 (52.5) | 236 (54.4) |
| Positive | 236 (35.5) | 83 (35.9) | 73 (34.0) | 80 (36.5) | 153 (35.3) |
| Other | 21 (3.2) | 8 (3.5) | 8 (3.7) | 5 (2.3) | 13 (3.0) |
| Symptom onset to randomization | | | | | |
| No. of patients | 584 | 209 | 181 | 194 | 375 |
| Time from symptom onset to randomization, median, d (IQR) | 3.0 (2–5) | 3.0 (2–5) | 4.0 (2–5) | 3.0 (2–5) | 3.0 (2–5) |

| | | | | | |
|---|---|---|---|---|---|
| At least one risk factor for hospitalization[d] | 408 (61.4) | 142 (61.5) | 134 (62.3) | 132 (60.3) | 266 (61.3) |

Data are presented as no. (%) unless otherwise indicated. Percentages may not total 100 because of rounding.
[a]Race and ethnic group were reported by the patients.
[b]The body-mass index is the weight in kilograms divided by the square of the height in meters.
[c]Obesity is defined as a body-mass index of greater than 30.
[d]Risk factors for hospitalization include an age of more than 50 years, obesity, cardiovascular disease (including hypertension), chronic lung disease (including asthma), chronic metabolic disease (including diabetes), chronic kidney disease (including receipt of dialysis), chronic liver disease, and immunocompromised (immunosuppression or receipt of immunosuppressants).
Abbreviations: IQR, interquartile range; RT-PCR, reverse-transcriptase polymerase chain reaction; SD, standard deviation.

#### Table S4. Change from Baseline in Viral Load ($log_{10}$ copies/ml) to Day 7 in Patients With No or ≥1 Risk Factors for Hospitalization

| End point | Placebo | REGEN-COV 2400 mg | REGEN-COV 8000 mg | REGEN-COV combined |
|---|---|---|---|---|
| **Change from baseline in viral load ($log_{10}$ copies/ml) to day 7 (analysis group 2)** | | | | |
| **No risk factors for hospitalization due to COVID-19 (mFAS)** | | | | |
| No. of patients | 55 | 48 | 58 | 106 |
| Least-squares mean change, $log_{10}$ copies/mL (SE) | −2.50 (0.21) | −2.90 (0.23) | −3.02 (0.21) | −2.97 (0.16) |
| 95% CI | −2.92, −2.08 | −3.34, −2.45 | −3.44, −2.60 | −3.28, −2.65 |
| Difference v. placebo at day 7 — $log_{10}$ copies/mL | | | | |
| Least-squares mean (SE) | | −0.39 (0.30) | −0.51 (0.29) | −0.47 (0.25) |
| 95% CI (*P* value)[a] | | −0.99, 0.20 (0.1933) | −1.08, 0.05 (0.0757) | −0.97, 0.03 (0.0677) |
| **≥1 risk factor for hospitalization due to COVID-19 (mFAS)** | | | | |
| No. of patients | 84 | 83 | 74 | 157 |
| Least-squares mean change, $log_{10}$ copies/mL (SE) | −2.56 (0.17) | −2.74 (0.17) | −3.08 (0.17) | −2.90 (0.13) |
| 95% CI | −2.90, −2.23 | −3.08, −2.41 | −3.42, −2.73 | −3.14, −2.65 |
| Difference vs placebo at day 7 — $log_{10}$ copies/mL | | | | |
| Least-squares mean (SE) | | −0.18 (0.23) | −0.51 (0.24) | −0.34 (0.20) |
| 95% CI (*P* value)[a] | | −0.63, 0.27 (0.4310) | −0.98, −0.05 (0.0302) | −0.73, 0.06 (0.0951) |

[a]*P* values are based on MMRM model with terms for baseline, baseline serology status, country, treatment, visit, treatment-by-visit, treatment-by-base, and base-by-visit interaction as fixed effect and subject as random effect.
Abbreviations: CI, confidence interval; LS, least squares; mFAS, modified full analysis set; SE, standard error.

#### Table S5. Description of COVID-19–Related MAVs[a]

| | Age range | Sex | Ethnicity | Race | Risk factor(s) (Y/N) | Symptom duration prior to randomization | Baseline viral load ($log_{10}$ copies/ mL) | Treatment arm | Type of MAV | Study day | Viral load at or around the time of MAV, $log_{10}$ copies/mL (d) | Reason for MAV | Details |
|---|---|---|---|---|---|---|---|---|---|---|---|---|---|
| 1 | 18 – 44 | M | Not Hispanic or Latino | White | Y | 7 days | 7.08 | Placebo | Hospitalization | 2 | 7.80 (1) | Pneumonia | 6-day hospitalization, did not require supplemental oxygen, ICU care, or mechanical ventilation |
| 2 | 65 – 84 | F | Hispanic or Latino | Not reported | Y | 6 days | 4.04 | Placebo | Hospitalization | 3 | 4.64 (3) | Hypoxemia | 5-day hospitalization, received supplemental oxygen. No ICU care or mechanical ventilation required |
| 3 | 18 – 44 | M | Hispanic or Latino | White | Y | 2 days | 7.85 | Placebo | Hospitalization | 9 | 5.97 (9) | Respiratory failure | 6-day hospitalization, treated with albuterol and supplemental oxygen. Did not require ICU care or mechanical ventilation |
| 4 | ≥85 | F | Hispanic or Latino | White | Y | 3 days | 7.85 | Placebo | Hospitalization | 10 | 5.51 (7) | Worsening of COVID-19/acute respiratory failure | 15-day hospitalization, treated with steroids and antibiotics, received supplemental oxygen. No ICU care or mechanical ventilation required |
| 5 | 45 – 64 | F | Hispanic or Latino | White | Y | 1 day | 7.85 | Placebo | Hospitalization | 10 | 5.22 (5) | Pneumonia | No additional details at this time |
| 6 | 45 – 64 | F | Not Hispanic or Latino | White | Y | 4 days | 7.09 | Placebo | ER visit | 5 | 5.42 (3) | Vomiting/Abdominal Pain | No additional details at this time |
| 7 | 45 – 64 | F | Not Hispanic or Latino | White | Y | pending | 7.85 | Placebo | ER visit | 10 | 5.02 (9) | Fever/Shortness of breath | No additional details at this time |

| | | | | | | | | | | | | | |
|---|---|---|---|---|---|---|---|---|---|---|---|---|---|
| 8 | 18 – 44 | F | Not Hispanic or Latino | Asian American | Y | 5 days | 4.41 | Placebo | ER visit | 13 | 3.24 (13) | Tachycardia | Prescribed ibuprofen and given IV normal saline |
| 9 | 18 – 44 | F | Not Hispanic or Latino | White | Y | 6 days | 6.23 | Placebo | ER visit | 15 | 0 (15) | Shortness of breath | Prescribed fluticasone and albuterol |
| 10 | 18 – 44 | F | Not Hispanic or Latino | White | N | 1 day | 6.59 | Placebo | ER visit | 21 | 0 (18) | Prolonged COVID-19 symptoms/fever/intermittent chest pain | Prescribed doxycycline and albuterol |
| 11 | 65 – 84 | M | Not Hispanic or Latino | White | Y | 2 days | 7.03 | Placebo | Phys office/telemed | 4 | 5.23 (3) | Cough | Prescribed benzonatate, ibuprofen |
| 12 | 18 – 44 | M | Not Hispanic or Latino | White | N | 6 days | 5.88 | Placebo | Phys office/telemed | 5 | 3.36 (5) | Persistent cough | Prescribed azithromycin and acetaminophen |
| 13 | 18 – 44 | M | Hispanic or Latino | White | Y | 4 days | 6.55 | Placebo | Phys office/telemed | 7 | 3.49 (7) | Worsening cough | Prescribed ceftriaxone, azithromycin, and ambroxol |
| 14 | 45 – 64 | F | Hispanic or Latino | White | Y | 3 days | 4.7 | Placebo | Phys office/telemed | 7 | 3.53 (7) | Worsening cough | Prescribed ceftriaxone, azithromycin, and ambroxol |
| 15 | 45 – 64 | F | Hispanic or Latino | White | Y | pending | 7.85 | Placebo | Phys office/telemed | 10 | 0 (9) | Worsening of COVID-19 | Prescribed promethazine |
| 16 | 18 – 44 | F | Not Hispanic or Latino | White | Y | 7 days | 3.99 | Low dose | Hospitalization | 2 | 3.99 (1) | Shortness of breath/pneumonia | 2-day hospitalization, treated with antibiotics, remdesivir, tocilizumab, and steroids. Did not require supplemental oxygen, ICU care, or mechanical ventilation |
| 17 | 65 – 84 | F | Hispanic or Latino | White | Y | 2 days | 6.73 | Low dose | Hospitalization | 3 | 6.73 (1) | Pneumonia | 7-day hospitalization, received supplemental oxygen. No ICU care or mechanical ventilation required |
| 18 | 18 – 44 | M | Not Hispanic or Latino | White | N | 7 days | 6.29 | Low dose | ER visit | 5 | 4.14 (5) | Pneumonia | No additional details at this time |

| | | | | | | | | | | | | | |
|---|---|---|---|---|---|---|---|---|---|---|---|---|---|
| 19 | 18 – 44 | F | Not Hispanic or Latino | White | N | 2 days | 7.85 | Low dose | ER visit | 5 | 3.83 (5) | Shortness of breath | No additional details at this time |
| 20 | 45 – 64 | F | Hispanic or Latino | White | Y | 6 days | 4.57 | Low dose | Phys office/telemed | 4 | 3.61 (3) | Cough/shortness of breath | Prescribed budesonide, salbutamol, and promethazine |
| 21 | 18 – 44 | M | Not Hispanic or Latino | Vietnamese | N | 5 days | 4.97 | Low dose | Urgent care clinic | 5 | 4.70 (5) | Shortness of breath/dropping oxygen saturation | Prescribed cefdinir, azithromycin, and combivent |
| 22 | 18 – 44 | M | Hispanic or Latino | White | Y | pending | 5.71 | High dose | Hospitalization | 1 | 5.71 (1) | Shortness of breath/worsening of COVID-19 infection | 7-day hospitalization, required supplemental oxygen and ICU care. Did not require mechanical ventilation. |
| 23 | 18 – 44 | F | Not Hispanic or Latino | White | N | 4 days | 7.24 | High dose[b] | ER visit | 3 | 4.95 (3) | Dyspnea | No additional details at this time |
| 24 | 18 – 44 | F | Not Hispanic or Latino | White, Native American, or Alaska Native | Y | 2 days | 5.45 | High dose | ER visit | 2 | 5.45 (1) | Shortness of breath | Prescribed ketorolac, dexamethasone, and ondansetron |
| 25 | 45 – 64 | M | Not Hispanic or Latino | White | Y | 3 days | 7.85 | High dose | ER visit | 2 | 7.90 (1) | Chest pain | No additional details at this time |
| 26 | 18 – 44 | F | Not Hispanic or Latino | White | N | 1 day | 7.85 | High dose | Phys office/telemed | 8 | 4.11 (7) | Pleurisy | Prescribed prednisone |
| 27 | 45 – 64 | F | Not Hispanic or Latino | White | Y | 6 days | 4.84 | High dose | Urgent care clinic | 10 | 0 (9) | COVID-19 symptoms | Prescribed azithromycin and prednisone |

[a]Analysis groups 1+2; modified Full Analysis Set.
[b]Only received 12.7 mL of infusion, stopped due to possible infusion-related reaction.
Abbreviations: ER, emergency room; ICU, intensive care unit; MAV, medically attended visit.

#### Table S6. Key Clinical End Points (Full Analysis Set)

| End point | Placebo | REGEN-COV 2400 mg | REGEN-COV 8000 mg | REGEN-COV combined |
|---|---|---|---|---|
| **Proportion of patients with COVID-19–related MAVs through day 29 (analysis groups 1+2)** | | | | |
| **FAS** | | | | |
| No. of patients | 266 | 266 | 267 | 533 |
| Patients with ≥1 visit within 29 days, no. (%) | 16 (6.0) | 8 (3.0) | 7 (2.6) | 15 (2.8) |
| Difference vs placebo | | −3.0% | −3.4% | −3.2% |
| **Proportion of patients with one of a subset of COVID-19–related MAVs consisting of hospitalization, emergency room visit, or urgent care visit through day 29 (analysis groups 1+2)** | | | | |
| **FAS** | | | | |
| No. of patients | 266 | 266 | 267 | 533 |
| Patients with ≥1 visit within 29 days, (%) | 10 (3.8) | 6 (2.3) | 6 (2.2) | 12 (2.3) |
| Difference vs placebo | | −1.5% | −1.5% | −1.5% |

Abbreviations: FAS, full analysis set; MAV, medically attended visit.

#### Table S7. Proportion of Patients Who Were Hospitalized, Visited the ER, and/or Died

| End point | Placebo | REGEN-COV 2400 mg | REGEN-COV 8000 mg | REGEN-COV combined |
|---|---|---|---|---|
| **Proportion of patients who were hospitalized or visited the ER through day 29 (analysis groups 1+2)** | | | | |
| **mFAS** | | | | |
| No. of patients | 231 | 215 | 219 | 434 |
| Patients with event within 29 days, no. (%) | 10 (4.3) | 4 (1.9) | 4 (1.8) | 8 (1.8) |
| Difference vs placebo | | −2.5% | -2.5% | -2.5% |
| 95% CI (*P* value)[a] | | −11.7, 6.8 (0.1766) | −11.7, 6.8 (0.1749) | −10.4, 5.5 (0.0778) |
| **Proportion of patients who were hospitalized or died through day 29 (analysis groups 1+2)** | | | | |
| **mFAS** | | | | |
| No. of patients | 231 | 215 | 219 | 434 |
| Patients with event within 29 days, no. (%) | 5 (2.2) | 2 (0.9) | 1 (0.5) | 3 (0.7) |
| Difference vs placebo | | −1.2% | −1.7% | −1.5% |
| 95% CI (*P* value)* | | −10.5, 8.0 (0.4516) | −10.9, 7.6 (0.2166) | −9.4, 6.5 (0.1339) |
| **Proportion of patients who were hospitalized through day 29 (analysis groups 1+2)** | | | | |
| **mFAS** | | | | |
| No. of patients | 231 | 215 | 219 | 434 |
| Patients with event within 29 days, no. (%) | 5 (2.2) | 2 (0.9) | 1 (0.5) | 3 (0.7) |
| Difference vs placebo | | −1.2% | −1.7% | −1.5% |
| 95% CI (*P* value)* | | −10.5, 8.0 (0.4516) | −10.9, 7.6 (0.2166) | −9.4, 6.5 (0.1339) |

[a]95% CIs and *P* values are based on exact method.
Abbreviations: CI, confidence interval; mFAS, modified full analysis set.

#### Table S8. Proportion of Patients With ≥1 COVID-19–Related MAVs by Baseline Serum Antibody Status

| End point | Placebo | REGEN-COV 2400 mg | REGEN-COV 8000 mg | REGEN-COV combined |
|---|---|---|---|---|
| **Proportion of patients with MAVs[a] through day 29 (analysis groups 1+2)** | | | | |
| **Baseline serum antibody status: negative (mFAS)** | | | | |
| No. of patients | 124 | 121 | 115 | 236 |
| Patients with ≥1 visit within 29 days, no. (%) | 12 (9.7) | 4 (3.3) | 4 (3.5) | 8 (3.4) |
| Difference vs placebo | | −6.4% | −6.2% | −6.3% |
| 95% CI (*P* value)[b] | | −13.4, −0.1 (0.0677) | −13.2, 0.3 (0.0704) | −13.2, −0.8 (0.0264) |
| **Baseline serum antibody status: positive (mFAS)** | | | | |
| No. of patients | 83 | 73 | 80 | 153 |
| Patients with ≥1 visit within 29 days, no. (%) | 2 (2.4) | 2 (2.7) | 1 (1.3) | 3 (2.0) |
| Difference vs placebo | | 0.3% | −1.2% | −0.4% |
| 95% CI (*P* value)[b] | | −6.1, 7.4 (1.0000) | −7.4, 4.7 (1.0000) | −6.8, 4.0 (1.0000) |
| **Baseline serum antibody status: other (mFAS)** | | | | |
| No. of patients | 24 | 21 | 24 | 45 |
| Patients with ≥1 visit within 29 days, no. (%) | 1 (4.2) | 0 | 1 (4.2) | 1 (2.2) |
| Difference vs placebo | | −4.2% | 0% | −1.9% |
| 95% CI (*P* value)[b] | | −21.1, 12.1 (1.0000) | −17.6, 17.6 (1.0000) | −19.4, 8.8 (1.0000) |

[a]COVID-19–related MAV included hospitalizations, ER visits, urgent care clinic visits, and outpatient/physician office/telemedicine visits.
[b]95% CIs and *P* values are based on exact method.
Abbreviations: CI, confidence interval; ER, emergency room; MAV, medically attended visit; mFAS, modified full analysis set.

#### Table S9. Proportion of Patients With ≥1 COVID-19–Related MAVs in Those With No or ≥1 Risk Factors for Hospitalization

| End point | Placebo | REGEN-COV 2400 mg | REGEN-COV 8000 mg | REGEN-COV combined |
|---|---|---|---|---|
| **Proportion of patients with COVID-19–related MAVs[a] through day 29 (analysis groups 1+2)** | | | | |
| **No risk factors for hospitalization due to COVID-19 (mFAS)** | | | | |
| No. of patients | 89 | 81 | 87 | 168 |
| Patients with ≥1 visit within 29 days, no. (%) | 2 (2.2) | 3 (3.7) | 2 (2.3) | 5 (3.0) |
| Difference vs placebo | | 1.5% | 0.1% | 0.7% |
| 95% CI (*P* value)[b] | | −4.7, 8.5 (0.6700) | −6.0, 6.3 (1.0000) | −5.3, 5.1 (1.0000) |
| **≥1 risk factor for hospitalization due to COVID-19 (mFAS)** | | | | |
| No. of patients | 142 | 134 | 132 | 266 |
| Patients with ≥1 visit within 29 days, no. (%) | 13 (9.2) | 3 (2.2) | 4 (3.0) | 7 (2.6) |
| Difference vs placebo | | −6.9% | −6.1% | −6.5% |
| 95% CI (*P* value)[b] | | −13.2, −1.3 (0.0185) | −12.6, −0.3 (0.0447) | −12.7, −1.6 (0.0065) |

[a]COVID-19–related MAVs included hospitalizations, ER visits, urgent care clinic visits, and outpatient/physician office/telemedicine visits.
[b]95% CIs and *P* values are based on exact method.
Abbreviations: CI, confidence interval; ER, emergency room; MAV, medically attended visit; mFAS, modified full analysis set.

#### Table S10. Proportion of Patients With ≥1 COVID-19–Related MAVs in Those Who Are Serum Antibody–Negative, High-Risk, and Have a Viral Load >$10^4$

| End Point | Placebo | REGEN-COV 2400 mg | REGEN-COV 8000 mg | REGEN-COV combined |
|---|---|---|---|---|
| **Proportion of patients with COVID-19–related MAVs[a] through day 29 (analysis groups 1+2)** | | | | |
| **Seronegative and viral load >$10^4$ copies/mL and high-risk** | | | | |
| No. of patients | 76 | 79 | 62 | 141 |
| Patients with ≥1 visit within 29 days, no. (%) | 10 (13.2) | 1 (1.3) | 2 (3.2) | 3 (2.1) |
| Difference vs placebo | | −11.9% | −9.9% | −11.0% |
| 95% CI (*P* value)[b] | | −22.0, −4.0 (0.0042) | −20.0, −0.1 (0.0651) | −20.8, −2.9 (0.0019) |

[a]COVID-19–related MAV included hospitalizations, ER visits, urgent care clinic visits, and outpatient/physician office/telemedicine visits.
[b]95% CIs and *P* values are based on exact method.
Abbreviations: CI, confidence interval; ER, emergency room; MAV, medically attended visit.

#### Table S11. Treatment-Emergent Serious Adverse Events and Adverse Events of Special Interest Reported in Subjects Receiving REGEN-COV

| System Organ Class<br>Preferred Term | Placebo Group (n = 262) | REGEN-COV 2400 mg (n = 258) | REGEN-COV 8000 mg (n = 260) | REGEN-COV combined (n = 518) |
|---|---|---|---|---|
| **Serious adverse events[a]** | | | | |
| **Gastrointestinal disorders** | | | | |
| Vomiting | 0 | 1 (0.4) | 0 | 1 (0.2) |
| Intestinal obstruction | 0 | 0 | 1 (0.4) | 1 (0.2) |
| Nausea | 0 | 1 (0.4) | 0 | 1 (0.2) |
| **Vascular disorders** | | | | |
| Hypertension | 1 (0.4) | 0 | 0 | 0 |
| **Respiratory, thoracic, and mediastinal disorders** | | | | |
| Hypoxia | 2 (0.8) | 0 | 0 | 0 |
| Dyspnea | 0 | 0 | 1 (0.4) | 1 (0.2) |
| **Metabolism and nutrition disorders** | | | | |
| Hyperglycemia | 0 | 1 (0.4) | 0 | 1 (0.2) |
| **Infections and infestations** | | | | |
| Pneumonia | 2 (0.8) | 1 (0.4) | 0 | 1 (0.2) |
| COVID-19 | 1 (0.4) | 0 | 0 | 0 |
| COVID-19 pneumonia | 0 | 1 (0.4) | 0 | 1 (0.2) |
| **Adverse events of special interest[a]** | | | | |
| **Gastrointestinal disorders** | | | | |
| Abdominal pain | 0 | 0 | 1 (0.4) | 1 (0.2) |
| Vomiting | 1 (0.4) | 0 | 0 | 0 |
| Nausea | 1 (0.4) | 0 | 0 | 0 |
| **Skin and subcutaneous tissue disorders** | | | | |
| Pruritus | 0 | 0 | 1 (0.4) | 1 (0.2) |
| Urticaria | 0 | 0 | 1 (0.4) | 1 (0.2) |
| Rash | 1 (0.4) | 0 | 0 | 0 |
| **General disorders and administration-site conditions** | | | | |
| Chills | 0 | 0 | 1 (0.4) | 1 (0.2) |
| Pyrexia | 0 | 0 | 1 (0.4) | 1 (0.2) |
| **Vascular disorders** | | | | |
| Flushing | 0 | 0 | 1 (0.4) | 1 (0.2) |
| **Nervous system disorders** | | | | |
| Dizziness | 1 (0.4) | 0 | 0 | 0 |
| Headache | 1 (0.4) | 0 | 0 | 0 |
| **Injury, poisoning, and procedural complications** | | | | |
| Infusion-related reaction | 0 | 0 | 1 (0.4) | 1 (0.2) |

Data are presented as no. (%) unless otherwise indicated.

[a]Only serious adverse events and adverse events of special interest (grade 2 or higher infusion-related reactions and hypersensitivity reactions) were collected.

#### Table S12. Mean Concentrations of REGN10933 and REGN10987 in Serum

| Nominal Sampling Time | REGN10933 (casirivimab) | | REGN10987 (imdevimab) | |
|---|---|---|---|---|
| | **1200 mg** | **4000 mg** | **1200 mg** | **4000 mg** |
| Predose | 0.0578 (0.594) [202] | 0.0421 (0.611) [210] | 0.0640 (0.603) [186] | 0 (0) [195] |
| End of Infusion[a] | 333 (86.8) [135] | 1022 (341) [126] | 336 (106) [136] | 1037 (332) [125] |
| Day 29[b] | 79.7 (34.6) [210] | 250 (97.4) [223] | 65.2 (28.1) [212] | 205 (82.7) [222] |

Data are presented as mean (SD) [N], where N is number of observations.
[a]Infusion duration was 1 hour.
[b]Observed concentration 28 days after dosing, i.e., on day 29.
Abbreviation: SD, standard deviation.
